# High-Quality and Easy-to-Regenerate Personal Filter

**DOI:** 10.1101/2021.12.16.21267766

**Authors:** Max Fraenkl, Milos Krbal, Jakub Houdek, Zuzana Olmrova Zmrhalova, Borivoj Prokes, Petr Hejda, Stanislav Slang, Jan Prikryl, Jakub Ondracek, Otakar Makes, Juraj Kostyk, Petr Nasadil, Pavel Malcik, Vladimir Zdimal, Miroslav Vlcek

## Abstract

Proper respiratory tract protection is the key factor to limiting the rate of COVID-19 spread and providing a safe environment for health care workers. Traditional N95 (FFP2) respirators are not easy to regenerate and thus create certain financial and ecological burdens; moreover, their quality may vary significantly. A solution that would overcome these disadvantages is desirable. In this study a commercially available knit polyester fleece fabric was selected as the filter material, and a total of 25 filters of different areas and thicknesses were prepared. Then, the size-resolved filtration efficiency (40–400 nm) and pressure drop were evaluated at a volumetric flow rate of 95 L/min. We showed the excellent synergistic effect of expanding the filtration area and increasing the number of filtering layers on the filtration efficiency; a filter cartridge with 8 layers of knit polyester fabric with a surface area of 900 cm^2^ and sized 25 × 14 × 8 cm achieved filtration efficiencies of 98 % at 95 L/min and 99.5 % at 30 L/min. The assembled filter kit consists of a filter cartridge (14 Pa) carried in a small backpack connected to a half mask with a total pressure drop of 84 Pa at 95 L/min. In addition, it is reusable, and the filter material can be regenerated at least ten times by simple methods, such as boiling. We have demonstrated a novel approach for creating high-quality and easy-to-breathe-through respiratory protective equipment that reduces operating costs and is a green solution because it is easy to regenerate.

## Introduction

The outbreak of COVID-19 caused by the SARS-CoV-2 virus and the subsequent explosive spread around the world have significantly increased the demand for highly efficient personal protective equipment, which is one of the most effective ways to reduce the exponential growth in the number of infected people.[1–3] Respiratory protection in particular is a decisive factor, as the SARS-CoV-2 virus is easily transmitted through the air by means of ballistic droplets larger than 100 μm that fall down very quickly and aerosol airborne particles that can easily travel in the air and accumulate in enclosed areas.[4–6]

While homemade cloth masks primarily protect others and can effectively absorb droplets produced by sneezing, coughing and speaking[3], they are inadequate for personal protection against smaller aerosol particles. In a hazardous environment, it is recommended to wear a filtering facepiece respirator (FFR) of at least class N95 or its European equivalent FFP2 or higher.[7]

Typically, the FFRs used derive their filtering properties from a very thin (0.1–1 mm) sandwich of fibrous layers. Improper filter layer manufacturing technology can result in low filtration efficiency or a high respirator pressure drop. Recent Canadian studies have shown large variations in filtration efficiency and pressure drop for N95, FFP2 and KN95 FFRs.[8,9]

At the time of FFR unavailability or for reasons of cost reduction, a suitable method of their regeneration can be used. The most common methods are: microwave-generated steam processing, ultraviolet germicidal irradiation, vaporized hydrogen peroxide. When choosing a method, it is necessary to take into account the type of regenerated respirator to avoid any damage or reduction in tightness and filtration efficiency. After regeneration, it is necessary to perform a thorough visual inspection and fit testing. Although correctly chosen methods are applicable for the regeneration of FFRs, their use is limited due to operating and acquisition costs, especially in LMICs[10–12]

On the other hand, since the outbreak of the Covid-19 crisis, many alternative fabric materials available for the manufacture of filter cloths have been explored.[13,14] The filtration efficiency of fabric materials was mostly below 30%.[15] To improve the filtration efficiency, the number of layers has been increased, but this inevitably leads to an increase in the pressure drop and the risk of unfiltered air being sucked in by leaks. However, a significant advantage of using certain textile materials is that they can be easily regenerated by exposure to elevated temperatures. Exposure of the fabric material to a temperature above 90 °C for 10 minutes is sufficient to inactivate the SARS-CoV-2 virus.[16,17] Inactivation of the virus can easily be achieved by boiling in water for the stated time or by washing in a washing machine at a temperature of 95 °C. This type of regeneration removes dirt, dust and SARS-CoV-2 virus.

Thus, the disadvantages of both methods of respiratory protection, i.e. the use of typical respirators and/or alternative face masks, inevitably provides an opportunity for a solution that at the same time achieves very good filtration efficiency as well as low respiratory resistance and is easy to regenerate. Such a solution will reduce the amount of waste produced and make available high-quality respiratory protection even where financial resources are limited.[18– 20]. (ref).

In this work, we have shown an excellent synergistic effect of expanding the filtration area and increasing the number of filtering layers on filtration efficiency. Using this approach, a filtration efficiency above 99% can easily be achieved at a volume flow rate of 95 L/min while maintaining low breath resistance. As the filter material we used a commonly available polyester knitted fleece which shows high mechanical and thermal stability. It can therefore be regenerated in very simple ways, such as boiling in water or washing at 95 °C. Based on the above, we designed and tested a personal filter prototype.

## Materials and Methods

In the present study, two common materials were tested as filtering materials: (1) single-faced jersey knit fabric, double-sided fleece, 100% polyester (PES), with a thickness of 2.3 mm and mass per unit area of 163 g.m^-2^, and (2) knit fabric, 100% cotton, with a thickness of 0.6 mm and area density 152 g.m^-2^ (Table S1 and Figure S1). Both materials were washed once at 95 °C before testing. The accessories for our designed filter kit consist of two flexible hoses for inhalers with an inner diameter of 22 mm and a length of 0.5 m manufactured by Technologie Médicale, Matala FSM-365 highly aerobic 3-dimensional structure, and a 3M 7500 series half mask.

To optimize the size and filtration efficiency of the filter, we prepared 25 filters with different filtering areas and filter thicknesses. The filter size was modified in multiples of the mask size 10 × 15 cm (150 cm^2^), and the filter thickness was increased by increasing the number of PES fabric layers (Figure S2).

The size-resolved filtration efficiency of the prepared filters was evaluated in the range of 40-400 nm with a scanning mobility particle sizer (SMPS) (3936L 75, TSI, USA). The volumetric flow rate was 95 L/min. A polydisperse challenging aerosol was generated by a nebulizer (AGK-2000 Palas) (Figure S3). The proposed filter cartridge was subsequently validated by the Czech Occupational Safety Research Institute according to the following standards: EN 143:2000/A1:2006

To determine the permeability of the used filtration materials, the pressure drop was measured on a setup consisting of a SC 15D scroll vacuum pump (Leybold), PG 07 and PG 08 flow meter (Rheotest), measuring chamber and G1107 manometer (Greisinger) with a resolution of 0.1 Pa (Figure S4).

The regeneration procedures included either boiling of the PES fabric for 10 minutes followed by drying, or washing in a washing machine at 95 °C with a washing powder and subsequent drying. Both procedures are suitable because they maintain the temperature above 90 °C for 10 minutes, which is sufficient to inactivate the SARS-CoV-2 virus and at the same time remove dust particles.[16,17]

## Results

In the present study, we propose two mutually supportive approaches that can increase the residence time of airborne particles inside a filter as they pass through, thereby improving the filtration efficiency of ultrafine particles. Both approaches are schematically illustrated in Figure 1, which shows that the enlarged filter area significantly reduces the face velocity, prolonging the time in which particles can be trapped inside the filter (the diffusion mechanism of capture is enhanced).[21] Additionally, the number of penetrating particles decreases exponentially with an increasing number of filter layers.

**Fig 1.**
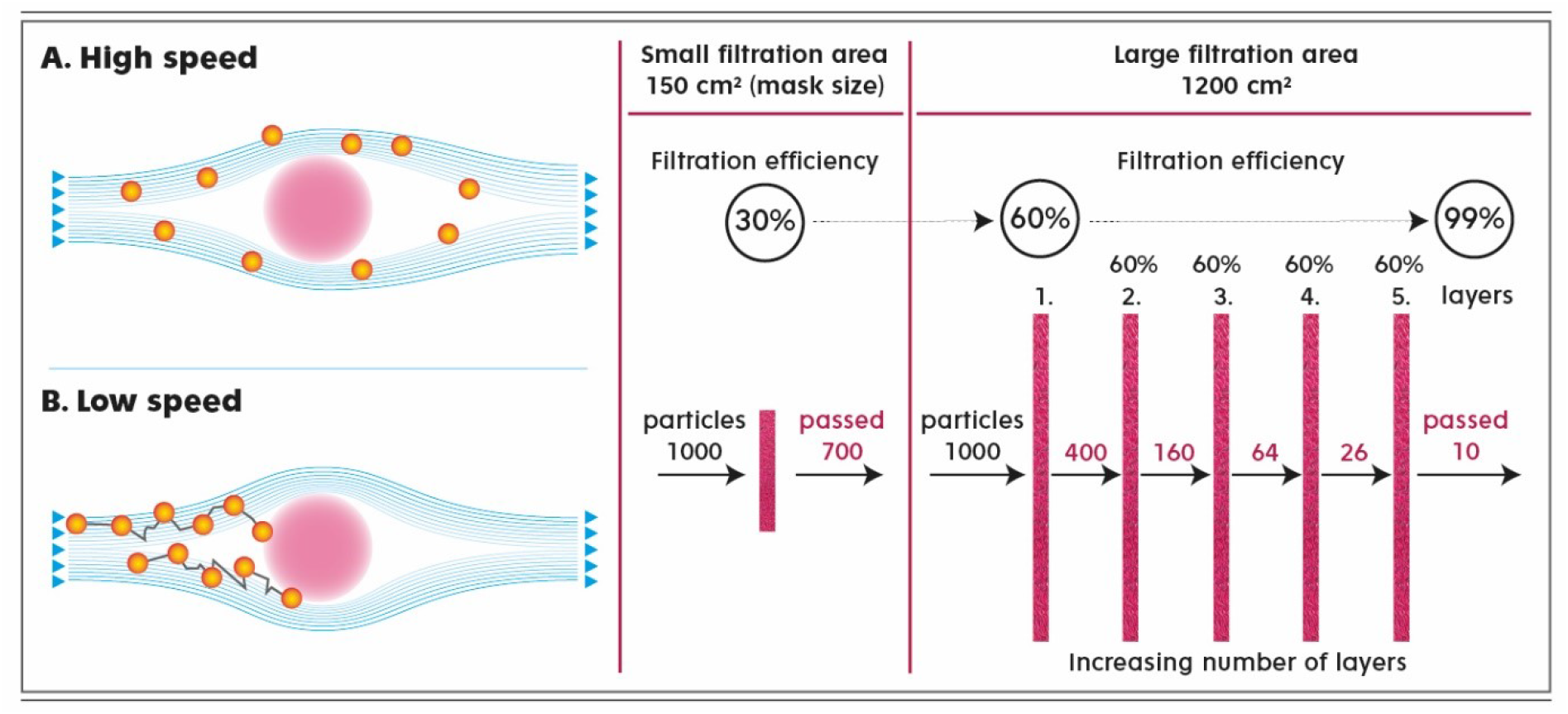
Principle of filtration efficiency enhancement. Panel A shows how the trajectory of the particle motion changes if we reduce the speed of the air that carries it around the filter fiber (cross-section). At low speeds (face velocities), as a result of Brownian motion, the particle deviates more from its original streamline, and the probability of it being trapped on the fiber surface increases (typically for particles smaller than 1 micron). Reducing the flow rate and thus increasing the filtration efficiency can be achieved by increasing the filter area, as shown in the example in Panel B. Increasing the number of filter layers leads to an exponential decrease in the number of particles passing through the filter and an increase in the filtration efficiency to 99%.

The filtration efficiency was measured for once washed (95 °C) PES fabric at a volumetric flow rate of 95 L/min corresponding to heavy physical work, as declared in EN 149:2001/A1:2009. The experimentally obtained values of filtration efficiency for a particle size of 100 nm depending on the size of the filter area and the number of layers of the PES fabric are summarized in Figure 2 (A). Here, we show that one layer of the PES fabric of the size of a standard mask shows a filtration efficiency of 30.5 %. The filtration efficiency further increases with increasing filtration area and number of PES fabric layers, with a maximum value of 99.1 % represented by 8 layers and 8 equivalent areas of a standard mask.

**Fig 2.**
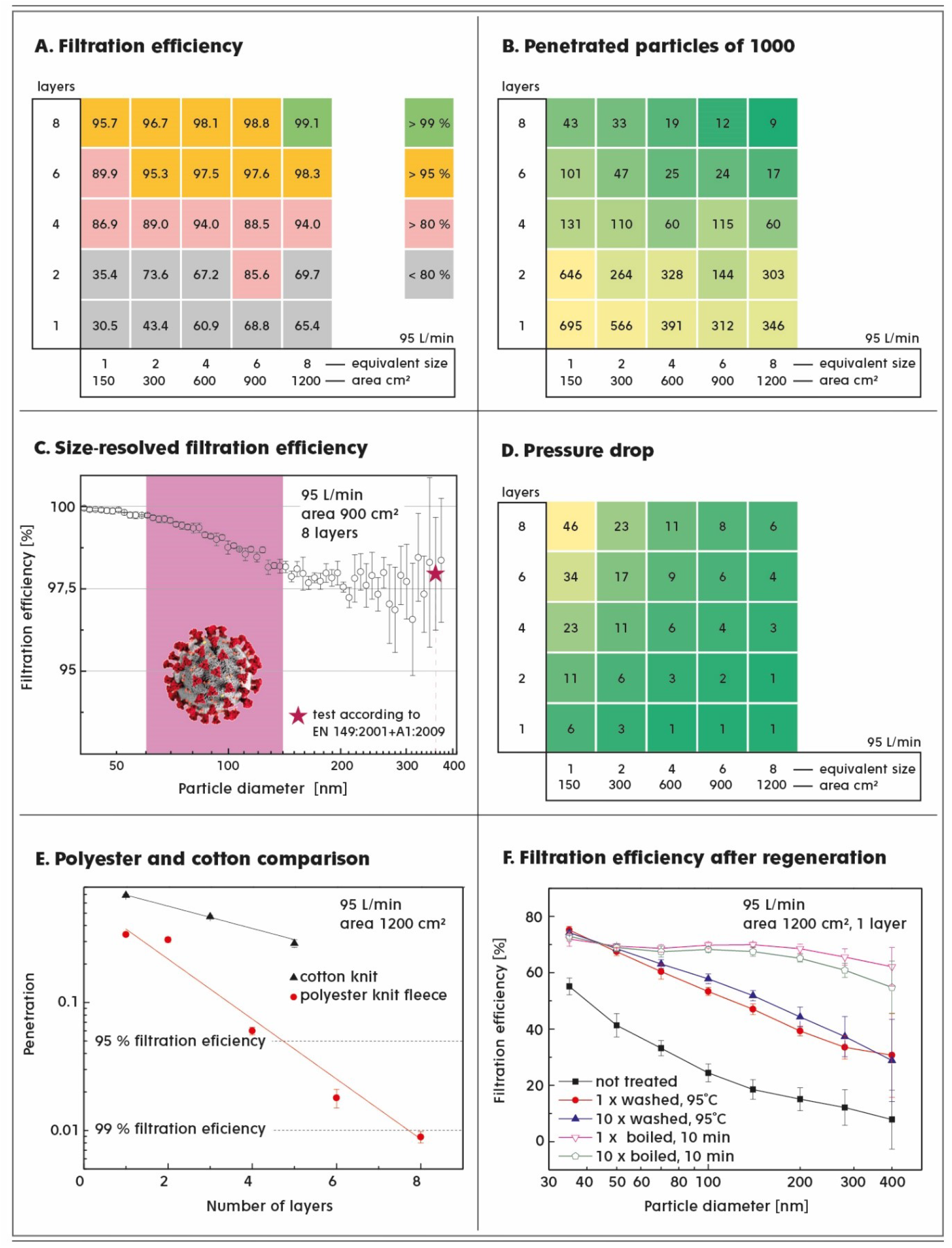
Characteristics of the knit PES fleece fabric used as a filter material. **Panels A, B and D** show the filtration ability of the PES fabric depending on the filter area, which was varied as multiples of the area of 150 cm^2^ (the size of the mask) and the number of layers of filter material. One layer has a thickness of 2.3 mm. The volumetric flow rate was selected to be 95 L/min. **Panel A** shows the filtration efficiency for 100 nm particles, **Panel B** represents the number of particles out of 1,000 that pass through the filter cartridge, and **Panel D** shows the corresponding pressure drop (Pa) evaluated on the basis of the measured permeability. **Panel C** shows the experimental curve of the filtration efficiency as a function of the particle diameter for a filter six times the size of a standard mask with eight layers of PES fabric. The blue star represents a measurement of the filtering efficiency performed by an accredited laboratory. **Panel E** demonstrates a comparison of the filtration efficiencies of two commonly available knit textile materials, namely, PES fleece and cotton, measured for 100 nm particles. **Panel F** shows the filtration ability of the PES fabric before and after washing at 95°C or boiling cycles.

However, particle penetration, rather than filtration efficiency, better describes the ability of the filter to protect the respiratory system. As shown in Figure 2B, the number of particles out of 1,000 that pass through the filter significantly differentiates between the quality of filters with very similar filtration efficiency. For instance, it is obvious from Figure 2B that the number of penetrating particles decreases more significantly with an increasing number of layers (bottom-up) than with increasing filter area (left – right). For the border cases, the best filter concept allows nine particles out of 1,000 to penetrate, which is approximately 70 times less than a single layer of a PES fabric filter with the size of a standard mask (150 cm^2^).

The filtration efficiency is affected by the size of the airborne particles, with a typical curve spanning particle sizes up to 400 nm, as shown for filter size 6 with 8 PES layers in Figure 2C and Figure S6. For small particles with a cutoff size of 80 nm, the diffusion filtration mechanism ensures better than 99% capture. The consequent reduction in filtration efficiency is the result of a gradual change in the filtration mechanism from exclusive diffusion to interception and impaction, reaching a minimum of 98 % for a particle diameter of approximately 300 nm.

Comfortable and easy breathing that does not cause considerable physical fatigue is one of the main prerequisites for the long-term use of RPE. Indeed, this aspect is associated with a pressure drop as a result of the filter’s resistance to the air flow. It is clear from Figure 2D that the assumption of expanding the size of the filter clearly reduces the pressure drop, while layering causes the opposite effect. From a practical point of view, it is necessary to take into account that one layer of a standard mask size has the same pressure drop as the 8-layer filter of 8 equivalent standard mask sizes, but the former shows 70 times higher penetration. Therefore, our results unambiguously represent the synergistic effect of the combination of layering and the expansion of the area of the filter and its possible applicability in filtration technology.

In this study, we compared PES and cotton fabrics, both knit, with similar fiber diameters and similar area densities (Table S1). Unlike the cotton fabric, the PES fabric had an additional fleece finish. As shown in Figure 2E, the particle penetration through the PES fabric was significantly lower than that through cotton fabric of the same filter area, demonstrating the filtration efficiency advantage of the additional fleece treatment (Figure S1).

Simple regeneration that can be repeated several times is a prerequisite for ecological RPE with low operating costs. It is worth exploring whether a simple treatment of the PES fabric by washing at 95 °C and boiling can impact the filtration properties of the selected material. Figure 2F shows that both procedures simulating the regeneration of the filter cartridge not only do not impair the filtration efficiency but also significantly improve the capture of airborne particles, especially after the first regeneration cycle, while subsequent cycles no longer affect the filtration ability of the PES fabric. During the regeneration process by washing at 95 °C, samples of PES fabric were taken after the first, third, fifth and tenth washes and the pressure drop was measured. The change in pressure drop of PES fabric between the first and subsequent wash cycles was less than 2%.

### The Filtration Kit

Using the results in Figure 2, we designed a filter cartridge composed of commonly available materials that is an inexpensive and easy-to-prepare alternative to commercial respirators. The supporting core of the filter cartridge was made on a 3D printer in the shape of a cuboid frame (21 × 14 × 4 cm), which alternatively can be replaced with a wooden or cardboard frame with an opening on one side for the insertion of a rubber hose.

The frame is further wrapped 8 times with the PES fabric to obtain a filter area of 900 cm^2^. The protruding PES fabric is firmly tightened with a twine at both ends to prevent ambient air from entering through these ends of the filter. For wearability, the filter prepared in this way is subsequently inserted into a small backpack that has two suction openings with a diameter of 4 cm at the bottom. To prevent contact between the filter and the walls of the backpack, porous plates are placed around the filter; these plates have an additional important function as air distributors. The air intake is located at the bottom of the filter carrier behind the user’s back, and the air flow in the entire kit is unidirectional, ensuring a constant supply of clean and oxygenated air. A detailed step-by-step assembly of the filter unit and a cross-sectional illustration of the filled backpack are shown in Figure 3 and in the Supplement.

**Fig 3.**
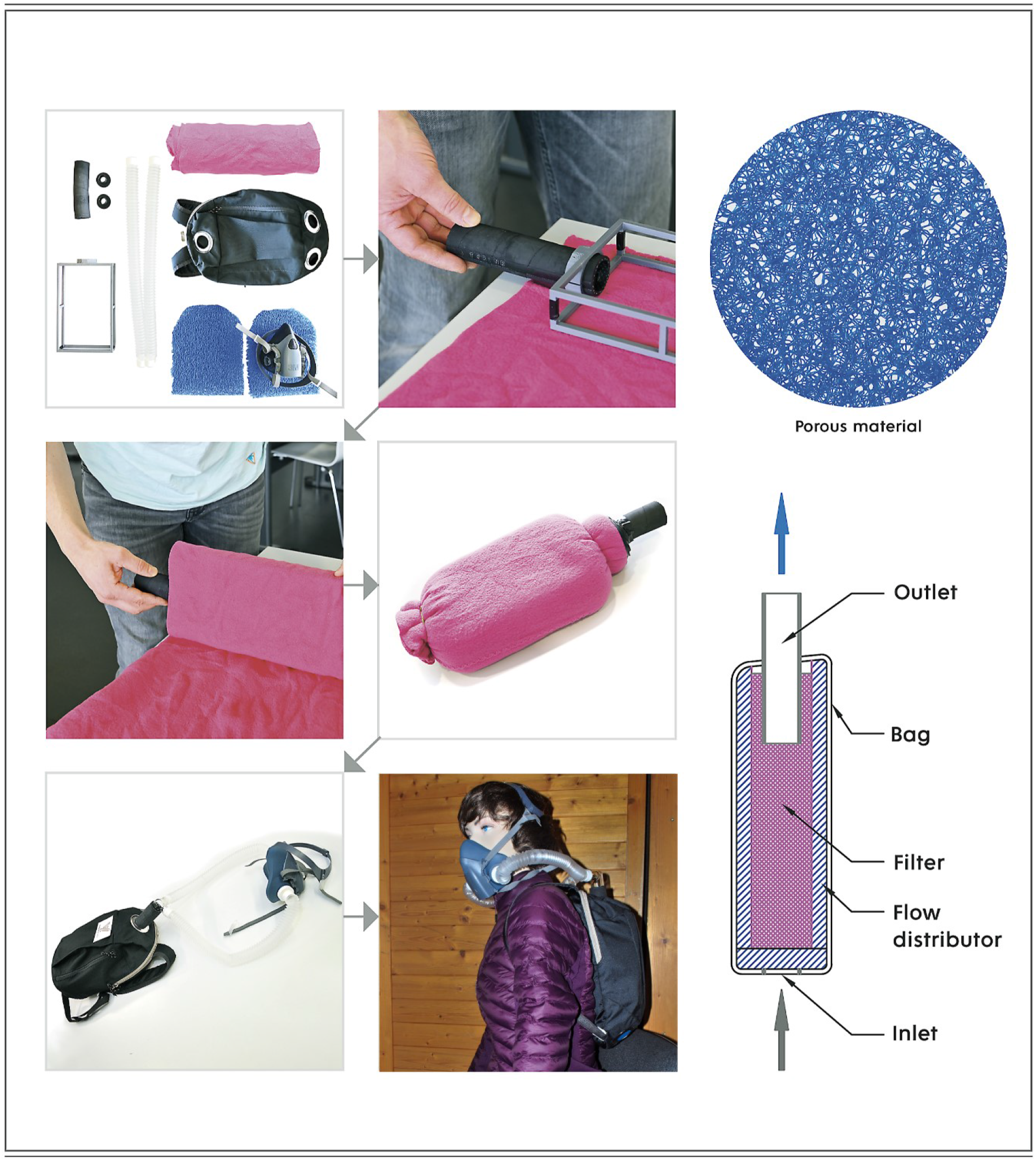
Assembly scheme of the filter kit. Shown are all parts of the filter kit and its gradual assembly, including a demonstration of how it is to be worn (manikin). The purpose of the blue porous material is to allow air to access the filter cartridge; air is sucked in over its entire surface. At the bottom right corner, the cross-section of the filter kit and the direction of air movement are shown.

In addition, the abovementioned filter cartridge was tested according to the EN 143:2000/A1:2006 standard by the Czech Occupational Safety Research Institute using a challenging salt aerosol with a count median particle diameter of 360 nm. The obtained filtration efficiency of 98 % at a volumetric flow rate of 95 L/min is in good agreement with our in-house measurements, as shown in Figure 2C and Figure S7.

Note that the key factor is the pressure drop throughout the filter kit to suppress the undesirable flow of contaminants through the gaps created between the skin and the half mask. The maximum acceptable value for the pressure drop of FFP2 respirators at a volumetric flow rate of 95 L/min as determined in the EN 149:2001/A1:2009 standard is 240 Pa. In the present case study, the kit consists of a filter cartridge, flexible hoses and a half mask, as shown in Figure 4. Experimentally, the pressure drop of the whole respiratory kit is 84 Pa, which is approximately 3 times better than the maximum permitted value. Note that the filter cartridge itself generates a pressure drop of only 14 Pa (see Figure S8) and that the remaining value is associated with the necessary accessories, which still leaves room for further possible improvements.

**Fig 4.**
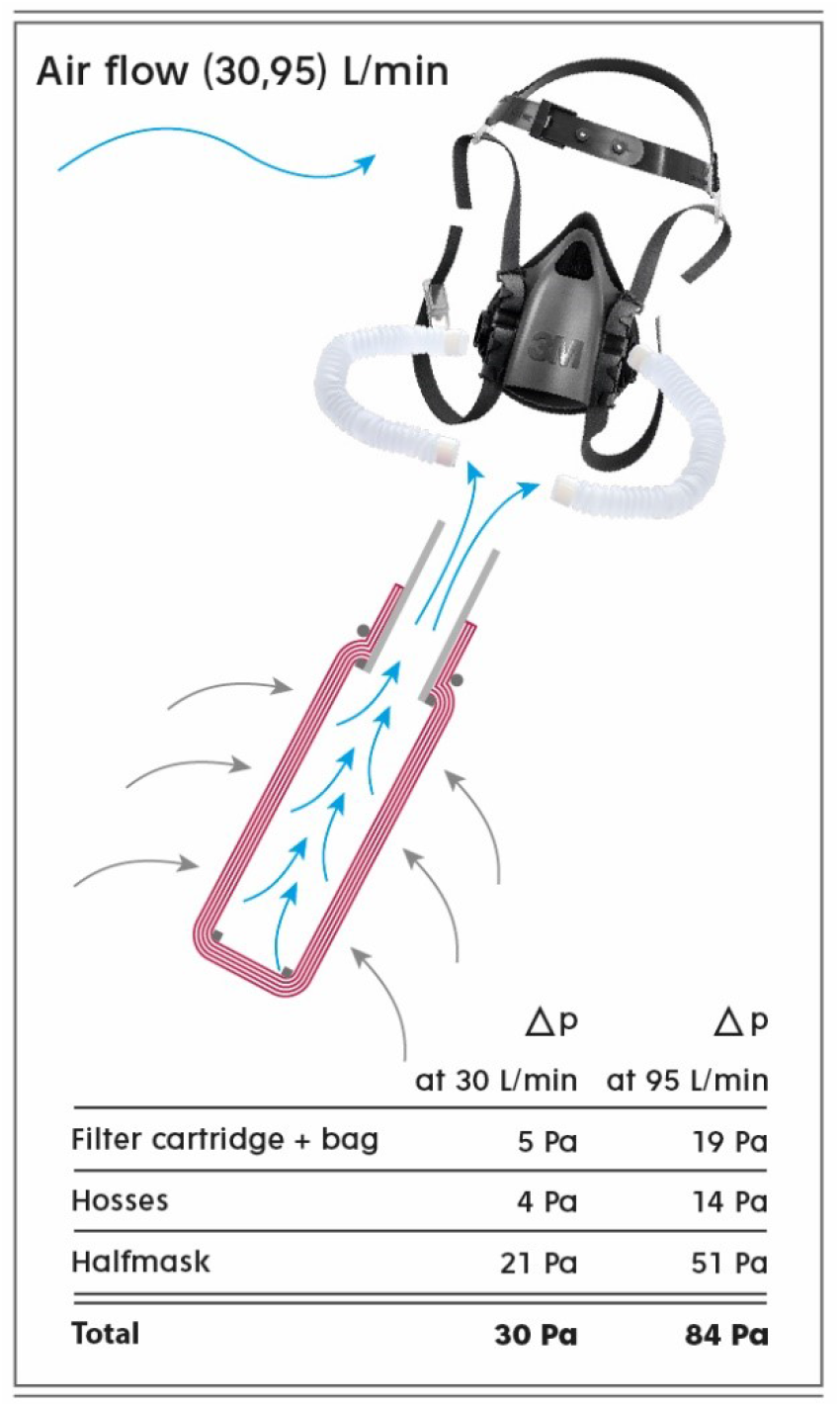
Pressure drop of the filter kit and its components. Shown are the pressure drop of the filter kit and its components at volumetric flow rates of 30 and 95 L/min, which correspond to air inhalation during light and heavy physical exercise, respectively. The cross-section of the filter cartridge is shown. The carrier (backpack) is not shown even though its pressure drop is taken into account.

## Discussion

### Filtering Efficiency and Pressure Drop

In the present work, we describe an RPE constructed from commonly available materials with a high filtration efficiency that meets the strict requirement for FFP2 (N95) respirators.[22]

A the size of a single virus particle (virion) has been estimated to be in the range of 60– 140 nm by electron microscopy analysis.[23] A recent study described the size distributions of airborne particles leaving an infected person who is speaking, singing, sneezing or coughing.[24] Particles that do not settle quickly decrease in size due to water evaporation and accumulate in the internal environment.[5,25] Therefore, we focused our filtration experiments on the border case represented by a particle size of 100 nm, as shown in Figure 2, while the results for particle diameters from 40 to 400 nm can be found in Figure S6 and Figure 2C.

Objectively, the strategies of increasing the filter surface and thickness are well known in filtration technology[21]; the combination of both approaches can be used in the preparation of a practical-size filter cartridge with a high filtration efficiency and low pressure drop. For example, a cuboid-shaped frame (21 × 14 × 4 cm) wrapped in 8 layers of PES fabric (Figure 3) has a minimum filtration efficiency of 98 % (99.5 %) and pressure drop of 14 (3) Pa at a volumetric flow rate of 95 (30) L/min, as demonstrated by the enclosed test report from the Czech Occupational Safety Research Institute (see Figures S7 and S8). The results obtained from the Czech Occupational Safety Research Institute agree well with in-house experiments (see Figure 2C). Note that the certified filter outperforms most of the 43 tested FFP2, N95 and KN95 class respirators, as published in recent studies[8,9] (see Figure S9). Naturally, by increasing the number of PES fabric layers to approximately 18 for the same filter size and particle diameters, the minimum filtration efficiency can reach 99.9 %, with a pressure drop of 29 Pa at a volumetric flow rate of 95 L/min (see Figures S10 and S11).

### Material Choice and Regeneration

The choice of a suitable fabric for filter fabrication is of undeniable importance. A few recent studies compared several household materials for homemade cloth face masks, and polypropylene, cotton and PES fabrics have been selected as the best candidates.[13,26] Nonetheless, two important aspects should be taken into account: 1) surface treatment and 2) thermal stability at 100 °C to facilitate very simple regeneration, for example, boiling.[16]

We showed that fabric surface treatment can have a decisive influence on the filtration properties. In our study, we examined knit fabrics made of cotton and PES with similar parameters except that the PES fabric had a fleece finish that increased the total thickness of one layer approximately 3-fold (Figure S1) while significantly increasing the active area for potential capture of hazardous particles. As shown in Figure 2E, particle penetration through the PES fabric with a fleece treatment is approximately one order of magnitude lower than that of only knit cotton, indicating that the outer nonwoven-like fleece part of the PES fabric very strongly promotes the filtration ability, whereas the inner knit part maintains good mechanical resistance and uniformity. Note that PES knit fleece fabric outperforms other conventional fabrics in terms of filtration efficiency and pressure drop (Figure S12).

Easy and repeatable regeneration of the filter cartridge is a prerequisite for safe multiple use. We chose both boiling and washing at 95 °C as simple regeneration procedures. For regeneration, the PES fabric is removed from the frame, boiled or washed, dried and rewound back onto the frame. Surprisingly, the PES fabric, unlike cotton, significantly increases the capture of airborne particles, especially after the first regeneration cycle, while subsequent cycles no longer affect the filtration ability of the PES fabric. We assume that the increase in filtration properties after first boiling or washing at 95 °C is due to a combination of an increase in the density of the fabric due to shrinkage by 4 % while maintaining the structure of the fleece treatment and possibly removing chemicals (fabric softener and oil) that remain inside the PES fabric after its industrial production. Therefore, we recommend as a first step before the assembly of the filter cartridge to boil the PES fabric to improve and unify its filtration properties. (Please note that all measurements were performed on a single washed 95 °C PES fabric).

### Filter Kit

The filter kit we designed consists of a filter cartridge, hoses and a half or full face mask and can potentially be applied in areas where high filtration efficiency, easy regeneration and low operating cost are required. Due to its excellent adjustable filtration properties and low pressure drop, health care workers are one of the target groups, especially in situations where medical persons wearing our filter kit with a filter cartridge on their back are near a source of droplets containing virus particles, such as in discussions with a patient, intubation, and ear, nose, and throat (ENT) and stomach examination.[27–29] We assume that a greater distance and shielding by the body of the filter user results in substantially a lower dose of inhaled virus, as schematically shown in Figure S13. In addition to its main advantages, our solution has certain disadvantages. These include, in particular, reduced audibility through the half mask and daily disinfection of all components except the filter cartridge.[30,31] The advantages and disadvantages of the designed filter cartridge/kit are clearly summarized in Table 1. We did not perform the fit testing on human subjects in the study, however standard professional half (or full face) masks are tested and ensure proper fit when wearing the filter kit.

**Table 1.**
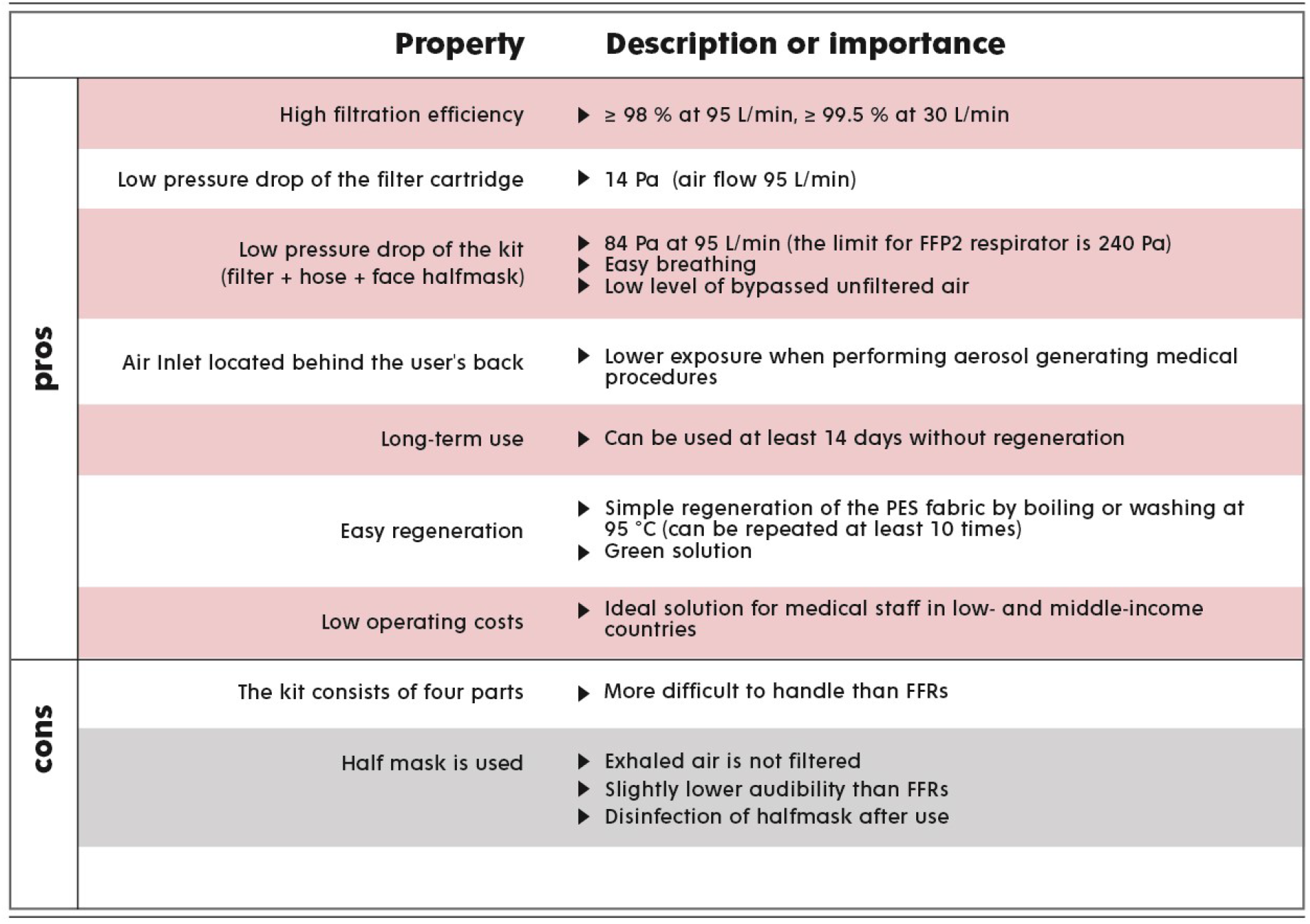
Summary of the advantages and disadvantages of the designed filter kit.

Finally, the acquisition and operating costs of the designed filter kit should be discussed. The filter cartridge inside the backpack can be used continuously without regeneration for at least 14 days, which corresponds to the use of P3 filters[30,32] (P3 filters are attached to a half or full face mask). Moreover, unlike the nanofiber membrane in the P3 filter, the PES material is less prone to clogging due to the larger pore size and thickness of the filter material.

With a simple calculation (10 regenerations) × (14 days of continuous filter use), it is possible to estimate the time that the PES fabric can be used as a filter material as up to 5 months, which is approximately equivalent to the use of 140 N95 or FFP2 standard FFRs with the consumption of 1 mask per day per person. We believe that the low operating costs, including mainly the cost of disinfection of the half mask and boiling water, make our filter kit the optimal solution for the proper protection of health care workers, especially in low- and middle-income countries. The total acquisition cost for the whole filtration kit is estimated at 56 EUR (retail price). The prices of individual components can be seen in Figure S14.

## Conclusion

We identified that the fleece treatment of the fabric has a positive effect on its filtration properties. Based on a detailed evaluation of the effect of the filter size and thickness on the pressure drop and filtration efficiency, we designed RPE where the filter cartridge is located on the user’s back. The proposed solution has a filtration efficiency easily tunable to values greater than 99.9% with a minimal increase in breath resistance; in addition, this solution better protects the user as he or she performs AGMPs than FFRs.

Because it can be easily regenerated, our proposed solution has the potential to reduce the environmental impact and simplify access to high-quality respiratory protective devices for health care workers, especially in low- and middle-income countries or in crisis situations.

## Data Availability

All data produced in the present work are contained in the manuscript

## Acknowledgment

This work was supported by the Ministry of Youth, Education and Sports of the Czech Republic (projects no. LM2018103 and LM201822).

We would like to thank prof. Oldrich Jirsak and Michal Cerny Ph.D. for technical advice, Ing. Dominik Stursa for technical support, prof. M.D. Roman Prymula for support in the early stage of the project to and M.D. Jan Vodicka for advice on the practical use of the personal filter.

## Notes

### Competing Interest Statement

The authors have declared no competing interest.

### Funding Statement

This study was funded by the Ministry of Youth, Education and Sports of the Czech Republic (projects no. LM2018103 and LM201822).

