## Supplementary_information for "High-Quality and Easy-to-Regenerate Personal Filter"

### Index

### Supplementary Methods

#### *1. Textile Materials Used*

Table S1

| sample | material | type of fabric | standard use | seller | areal density | thickness |
|---|---|---|---|---|---|---|
| | | | | | $g.m^{-2}$ | mm |
| A | 100% polyester | single-faced jersey fabric, double side fleece. | blanket | Vinnemeier GmbH | 163 | 2,3 |
| B | cotton | knitted fabric | T-shirt | VESNA | 152 | 0,6 |

| sample | material | fiber diameter | number of rows | number of columns | permeability |
|---|---|---|---|---|---|
| | | µm | per cm | per cm | $m^2$ |
| A | 100% polyester | 13.3 ± 0.1 | 12 | 12 | 8E-10 |
| B | cotton | 16.7 ± 2.7 | 20 | 15 | 3.7E-10 |

Test performed according normatives:

| | |
|---|---|
| areal density tested according | EN12127 |
| thickness | EN ISO 5084 |
| number of columns and rows | EN 14971 |

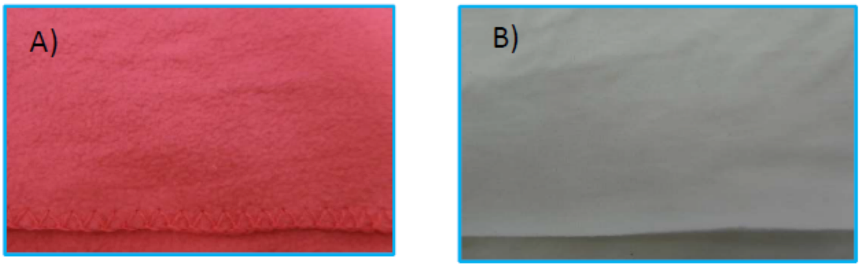

### 2. *Fleece Is a "2 in 1" Material*

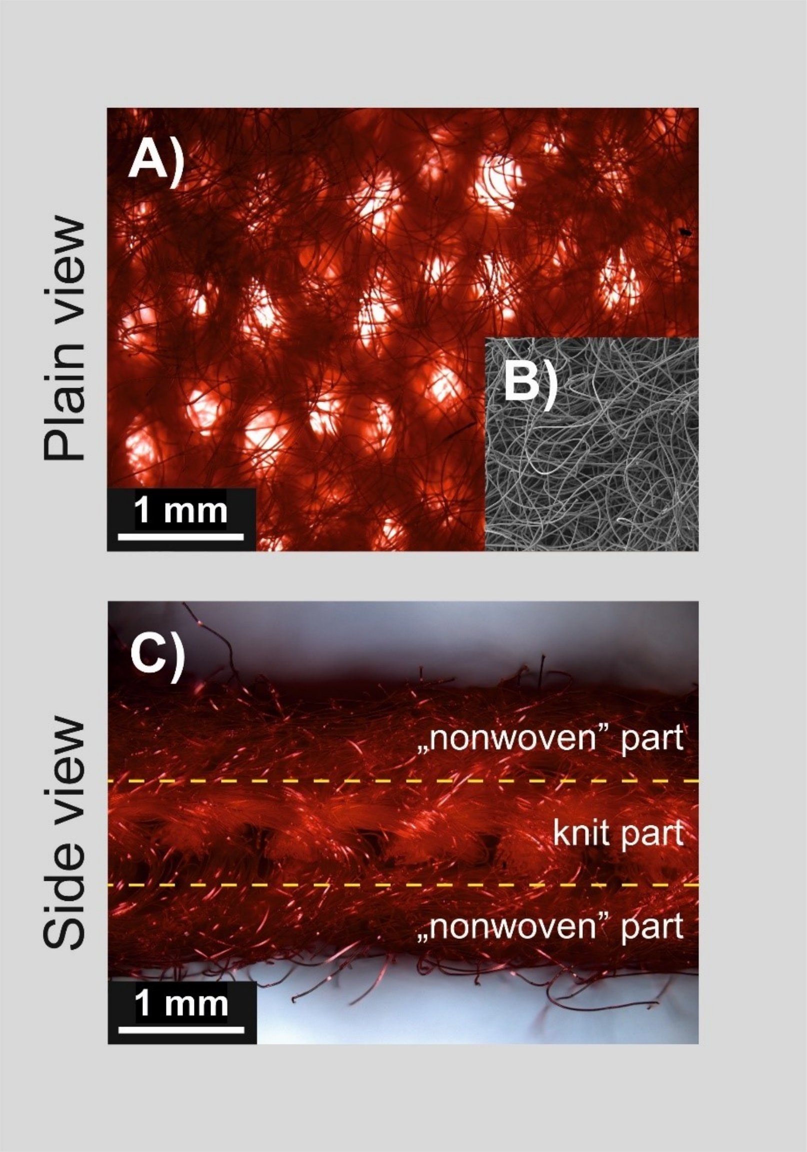

*Figure S1* Fleece is a soft napped fabric made of polyester. It is made from a brushed knit polyester fabric. Transmission optical photography allows us to see the knitted part of the fleece material (A), while scanning electron photography shows the outer part of fleece material, which remains a nonwoven filter layer (B). Both pictures A) and B) are of the same scale. In the side view, the two distinct parts of the fleece material can be recognized (C). Importantly, fleece material combines the good mechanical properties of knitted fabrics and the good filtering ability of a nonwoven filter layer.

### 3. Filter Cartridge Preparation for Measurement

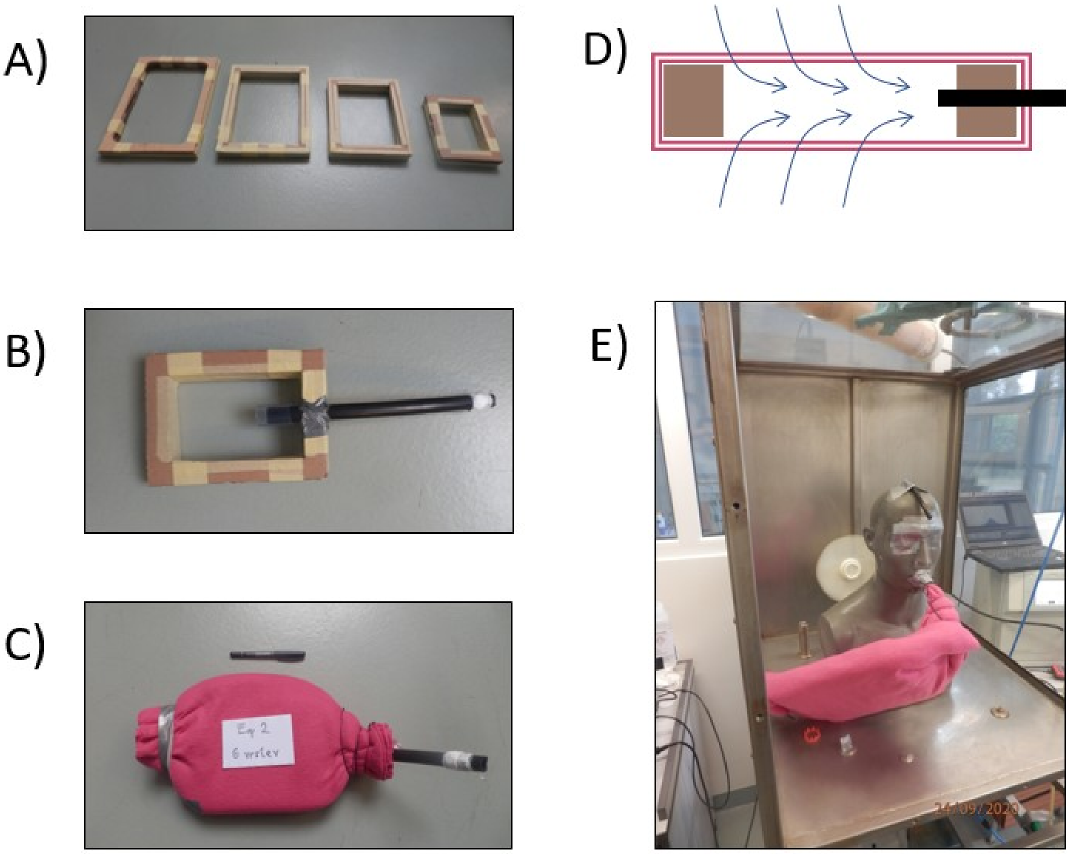

*Figure S2. The filter cartridge design for testing the filtration efficiency was based on different-size frames. Four of the frames are shown in panel (A). The filtering area values are 150, 300, 600, 900, and 1200 cm$^2$. The frame thickness is approximately 2 cm (see Table S2 and Table S3). The graphite rubber hose was tightened to the frame (B). Graphite suppresses aerosol charging, and thus, the measurement is not distorted. The frame prepared in this way was wrapped with an appropriate number of PES fleece layers (1, 2, 4, 6, or 8). The thickness of a single PES fleece layer was 2.3 mm. Both sides of the filter were tightened with twine. The frame filter (filter cartridge) allows air to pass via the upper and lower parts of the frame opening, and the filtered air passes inside the hose (D). The prepared filter cartridge was introduced in a measuring chamber, and the graphite hose was inserted in the manikin mouth and sealed (E). Then, the chamber was closed, and the filtration efficiency was measured.*

### *4. Size-resolved Penetration–Measurement Setup*

The penetration $P$ is complementary to the filtration efficiency $E$ *(%)*.

$$E = (1 - P) * 100 \qquad \text{(Eq. S1)}$$

The penetration represents the proportions of the particle concentrations behind and in front of the filter, while the filtration efficiency represents the percentage of particles trapped on the filter. In the case of respiratory protective equipment (RPE), we are interested in how many particles pass through the filter into our respiratory system, which is one of the advantages of working with results expressed as penetration values.

#### Measurement set-up

The size-resolved penetration through the material was measured using a filter testing system developed in cooperation with the Institute of Chemical Process Fundamentals of the Czech Academy of Sciences (ICPF CAS) and National Institute for Nuclear, Chemical and Biological Protection (NINCBP). A simplified diagram of the measurement apparatus is shown in Figure S3.

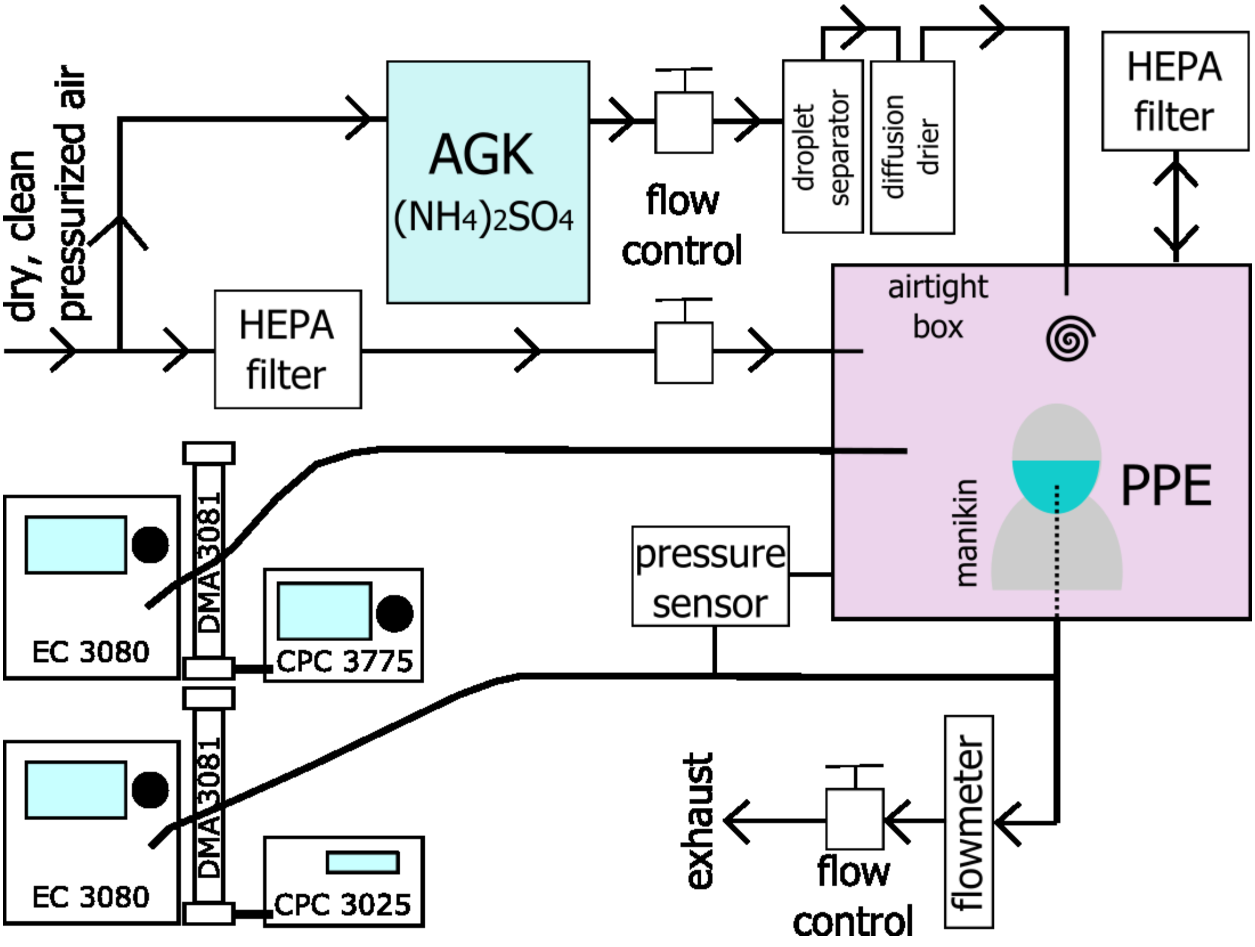

*Figure S3. Diagram of the measurement set-up. Dry clean pressurized air enters the AGK aerosol generator filled with $(NH_4)_2SO_4$ solution. The generated polydisperse aerosol continues through the droplet separator and diffusion driers into the airtight box, where the defined aerosol atmosphere is reached (the entering aerosol is mixed with dry clean air to achieve a homogeneous concentration in the whole box). The aerosol PNSD is measured using a set of two aerosol spectrometers, SMPSs, one sampling the aerosol in front and the other behind the RPE being tested. The measurement of the pressure drop across the RPE facilitates continuous monitoring of the possible changes in the respirator properties and at the same time provides information about the breathing resistance.*

The measurements were conducted at a volumetric flow rate of 95 L/min (with a corresponding face velocity of 10.6 cm/s), which corresponds to the more intense breathing activity of an adult during through personal protective equipment (respirator/face mask with a surface area of 150 $cm^2$) heavy physical exercise. In our case, the filtration efficiency of the filtering cartridge connected to the mouth of the manikin was tested. The challenging aerosol was generated by a nebulizer (AGK-2000, Palas) by dispersion of $(NH_4)_2SO_4$ salt solution (1 g/l). The operating conditions result in aerosol having geometric mean diameter (GMD) at approximately 70 nm with geometric standard deviation (GSD) around 1.6. The generated aerosol was passed through a diffusion dryer containing silica gel to be dried, then it was directed into an airtight box; at the entrance of this box, it was intensely mixed with dry particle-free air. The flow rate of dry air was controlled to sufficiently dilute the aerosol and to compensate for the outgoing amount of air being sampled through the head of the manikin.

Charge neutralizer was not used during these experiments. Due to long enough residence time in the Testing chamber (box) the testing aerosol should be most probably close to charge equilibria. The testing box was also connected to atmospheric pressure using a high-efficiency particulate absorbing (HEPA) filter to prevent the over/underpressurizing of the testing box. The tested RPE, in our case the filter cartridge, was inserted into the manikin mouth and sealed. The aerosol particle number size distribution (PNSD) was monitored during the whole measurement using a set of two aerosol spectrometers (scanning mobility particle sizers (SMPSs)). One spectrometer measured the particle size distribution outside the RPE (in the volume of the airtight box), while the other was sampled from the line taking the defined volumetric flow sucked in by the mouth of the manikin through the tested RPE (allowing us to take a sample behind/inside the tested RPE).

Before each measurement of the size-resolved filtration efficiency, the size-resolved correction factor between the two aerosol spectrometers was estimated to compensate for deviations between the two aerosol spectrometers. The changes in the filtering material properties during the measurement (e.g., loading by the challenging aerosol) were monitored by means of the measurement of the pressure drop across the filter material, which at the same time provided information about the breathing resistance. The description of the instruments used in the measurement setup is described in the following paragraphs.

### The instrumentation

#### Aerosol generator

A nebulizer (AGK-2000, Palas, Germany) was used as the source of a well-defined challenging aerosol. The aerosol was nebulized from $(NH_4)_2SO_4$ salt solution (1 g/l) using a particle-free dry air pressure of 2.5 bars. The resulting aerosol particles were in the range of approximately 40 to 400 nm. The count median diameter of the tested aerosol size distribution was close to 80 nm.

#### SMPS

The SMPS (3936NL, TSI, USA) facilitates the measurement of the number concentration of aerosol particles and their size distribution in the submicron size range (particle diameters < 1 μm). This spectrometer consists of two parts:

1) The **electrostatic classifier** (EC model 3080, TSI, USA) gradually selects individual size fractions (with a resolution of 64 size channels per decade) from the sampled originally polydisperse aerosol after it passes through an inertial impactor to remove supermicron particles and to define the upper boundary of the size distribution. Then, the dried polydisperse aerosol is brought to Boltzmann charge equilibrium during its passage through an aerosol neutralizer containing an $^{85}Kr$ source, and afterwards, the individual size fractions of aerosol particles are selected (one by one depending on the high voltage set on the inner electrode of the differential mobility analyzer) in the electrostatic field based on their electrical mobility.

2) The selected monodisperse aerosol fraction then passes to a **condensation particle counter** (CPC) (model 3775 and 3025, TSI, USA), where the number concentration of each fraction is measured. Inside the CPC, the individual particles increase in size because of the condensation of n-butanol on their surface, and the grown particles are then detected by an optical method (light scattering).

The PNSD is obtained after correction of the raw data for multiple charges, particle losses inside the spectrometer due to Brownian diffusion and axial dispersion, and the CPC counting efficiency and after data inversion based on transfer functions for individual size bins.

The zero-concentration test for both SMPS spectrometers was performed at the beginning of each measurement procedure. This test was based on clean air sampling from the testing box to confirm that the whole apparatus was airtight and that the measurement was not affected by any leaks. In a comparison of both SMPS spectrometers, the results showed a difference between the two spectrometers of less than 10 %. Nevertheless, to compensate for differences in their detection efficiency, a measurement without any RPE attached to the face of the manikin was performed to estimate the size-resolved correction factor between the two spectrometers prior to each measurement procedure.

#### Data treatment

The values of the number concentrations in the individual size bins of the PNSD upstream and downstream of the RPE were measured using two aerosol SMPS spectrometers. Several measurements (minimum 4) were conducted for each RPE to ensure statistically correct results. The median of the number concentration in each individual size bin was calculated for both aerosol spectrometers. Based on these values, the penetration through the tested RPE was evaluated for each size fraction ($P_i$) using the following equation:

$$P_i = \frac{c_{i,DOWN}}{c_{i,UP}} K_i \qquad \text{(Eq. S2)}$$

where $c_{i,DOWN}$ denotes the median number concentration in bin $i$ of a given size measured by aerosol spectrometer sampling downstream of the RPE, $c_{i,UP}$ represents the median number concentration in bin $i$ of a given size measured by aerosol spectrometer sampling upstream of the RPE, and $K_i$ stands for the correction factor between the two aerosol spectrometers for bin $i$ of a given size. The filtration efficiency for a given size bin ($E_i$) was based on the following equation:

$$E_i = (1 - P_i) \times 100 \qquad \textit{(Eq. S3)}$$

### 5. Filter Fabric Permeability Evaluation

The permeability of the filter material was evaluated to optimize the filter thickness and area. A higher permeability value leads to lower respiratory resistance. The evaluation of permeability $k$ ($m^2$) was performed according to Darcy's law

$$k = \frac{\mu Q L}{\Delta p A} \quad \text{(Eq. S4)}$$

based on the measurement of the pressure drop $\Delta p$ (Pa) when the air flow $Q$ ($m^3/s$) passes through a porous material of thickness $L$ (m) and area $A$ ($m^2$). The air properties are represented by the dynamic viscosity $\mu$ (Pa.s).

The measurement setup consisted of a measuring chamber with three cylindrical extensions (surfaces 40, 86, 182 $cm^2$), an SC 15D (Leybold) vacuum pump, two PG 07 and PG 08 (Rheotest) flow meters, and a G1107 (Greisinger) manometer with a resolution of 0.1 Pa (Figure S4).

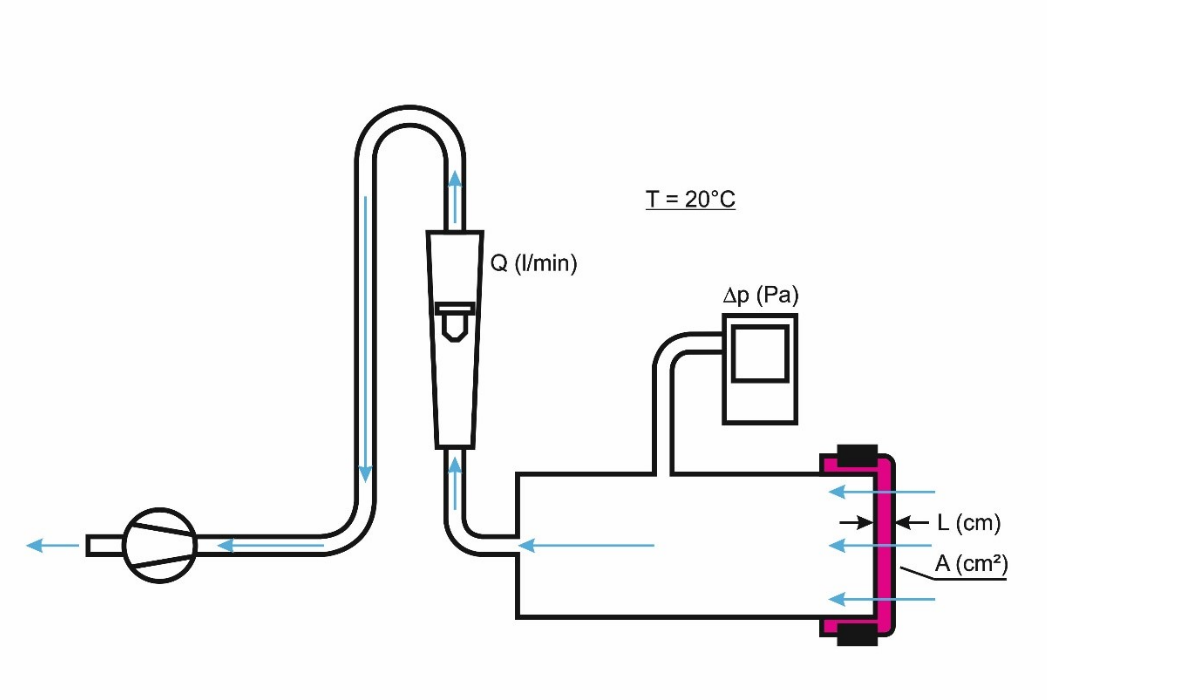

*Figure S4. The setup for measuring the fabric permeability consists of a measuring chamber to which the sample is attached, pressure gauge, flow meter and vacuum scroll pump.*

Measurements were made for 1 to 6 layers of filtering material, three different areas of the filtering material (40, 86, and 182 $cm^2$), and different air flow rates (10, 20, 30, 40, 60, 70, 80, 85, and 90 L/min). The corresponding air velocity ranged from 1 to 40 cm/s. The calculated permeability of the polyester knit fleece is shown in Figure S5. The picture shows the decreasing trend of the permeability value with increasing pressure drop. The average value of the permeability for the polyester (PES) fleece is $8\times10^{-10}$ $m^2 \pm 6$ %. The average value was taken for a pressure drop below 20 Pa; the filter was designed for use in this value range. The pressure drop of the polyester knit fleece fabric (Figure 2B) was calculated for a different filter area and thickness based on a known value of permeability (*(Eq. S4*). The permeability of the cotton textile is $3.7\times10^{-10}$ $m^2$. Under the same conditions, the polyester knit fleece fabric would have half the pressure drop of the cotton fabric.

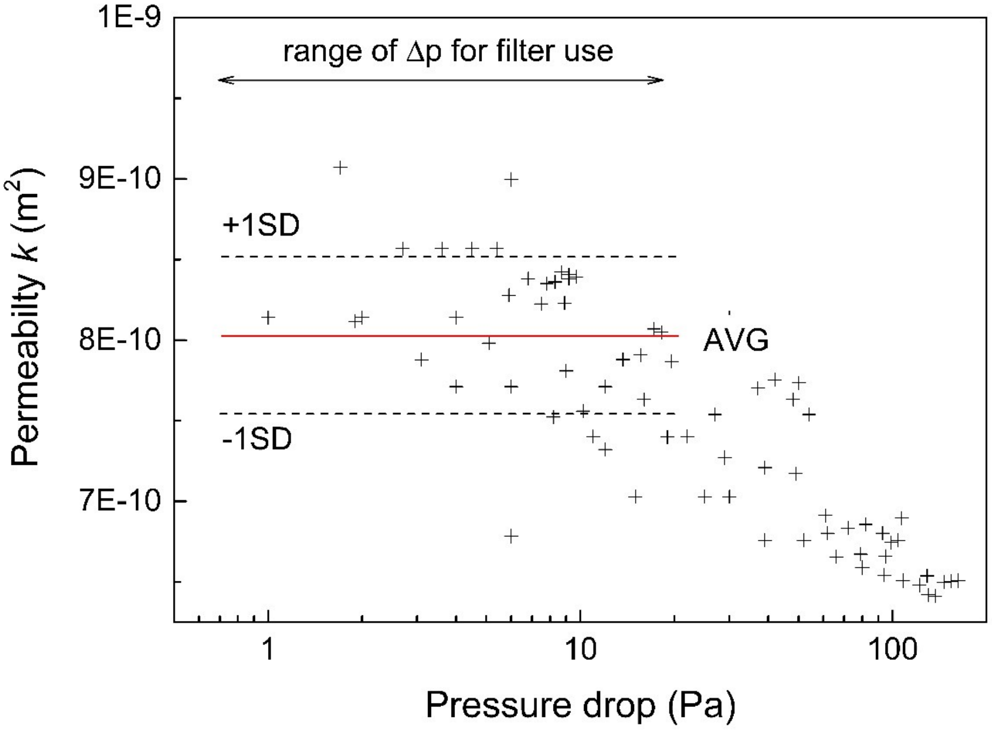

*Figure S5. Calculated permeability of the polyester knit fleece. Measurements of the pressure drop were made for different fabric thicknesses, areas and volumetric flow rates.*

### Supplementary Results

#### 6. Size-resolved Penetration Results

A set of 25 measurements of size-resolved penetration was performed (Figure S6). The filter area and thickness were changed accordingly (Table S2, Table S3)

*Table S2. Filter equivalent size. The filter size was modified in multiples of the mask size 10 × 15 cm (150 cm$^2$)*

| *Filter equivalent size* | *Filter area cm$^{-2}$* | *Face velocity cm.s$^{-1}$* |
|---|---|---|
| *1* | 150 | 10.6 |
| *2* | 300 | 5.3 |
| *4* | 600 | 2.6 |
| *6* | 900 | 1.8 |
| *8* | 1200 | 1.3 |

*Table S3. Filter thickness. The filter thickness was increased by the number of PES fabric layers*

| *Number of filter layers* | *Filter thickness (mm)* |
|---|---|
| *1* | 2.3 |
| *2* | 4.6 |
| *4* | 9.2 |
| *6* | 13.8 |
| *8* | 18.4 |

All of the measurements of the size-resolved penetration were performed at a volumetric flow rate of 95 L/min, which corresponds to the breathing rate of a person running or performing hard work. Each measurement was repeated 4 times, and the mean values are presented. The gradual decrease in the penetration values with increasing thickness (number of layers) is obvious in the range of particle sizes from 40 nm to 400 nm. The same applies to the filter area (size) according to Figures S6 (A-E). For easier interpretability of the filter performance, a dashed line is drawn. The results below the dashed line represent penetration lower than 0.05, which corresponds to a filtration efficiency higher than 95 %. An overlapping or crossing of curves is presented. These minor irregularities could be explained either by the filter preparation procedure or filter material (fleece) nonuniformity.

The improvement in the filtration efficiency (reduction in penetration) with increasing surface area for particles of all sizes (40-400 nm) can be explained in the first approximation as follows: reducing the face velocity increases the efficiency of the Brownian diffusion collection mechanism (for particles smaller than approximately 300 nm), while the interception collection mechanism is effective for particles larger than ca. 300 nm is independent of the face velocity.[1]

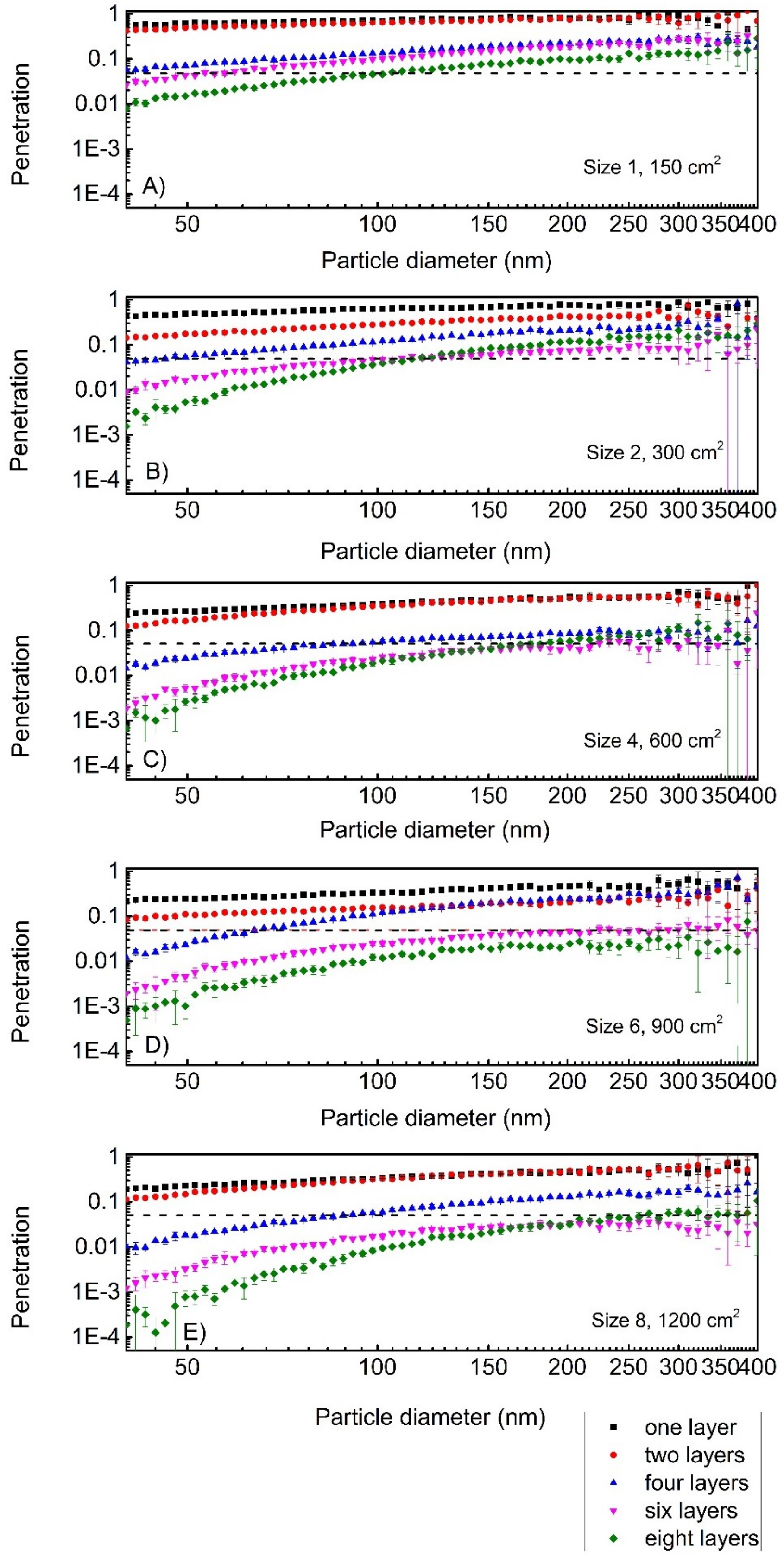

*Figure S6 shows the dependence between the penetration and particle diameter. The measurements were performed for the polyester knit fleece fabric at a volumetric flow rate of 95 L/min. The filter size increases from panel A to E. The color of the curve indicates the number of layers. The penetration values below the dashed line correspond to filtration efficiencies greater than 95 %.*

### *7. Report by an Accredited Laboratory*

Figure S7 shows a measurement report of the cartridge filtration efficiency (8 layers of PES fabric, surface 900 $cm^2$). The measurement was performed by the Czech Occupational and Safety Research Institute according to European standards for RPE testing EN149:2001+A1:2009. The measured penetration values of 2.07 % and 0.47 % correspond to filtration efficiency values of 98 % and 99.5 % for volumetric flow rates of 95 L/min and 30 L/min, respectively.

### Test report No. 054/2021

| | | | |
|---|---|---|---|
| Pages: | 3 | Prints-out: | 3 |
| Annexes: | 0 | Copy No.: | 1 |

| | |
|---|---|
| Client: | **Univerzita Pardubice, FCHT, CEMNAT, Studentská 95, 532 10 Pardubice, Czech Republic** |
| Test subject: | **PES filter cartridge 900 cm2** |
| Name of tests: | **Test according to EN 143** |
| Samples received: | 11. 2. 2021 |
| Test executed: | 12. 2. 2021 |
| Report issued : | 15. 2. 2021 |

Person authorized to sign the report:

Výzkumný ústav bezpečnosti práce, v. v. i. Akreditovaná zkušební laboratoř č. 1040 VÚBP Jeruzalémská 1283/9, 110 00 Praha 1

Ing. Lukáš Zavřel
*Head of VÚBP-ZL*

Distribution List:
1. Client
2. Laboratory Archive
3. VUBP-ZL Secretariat

Test results apply only to the tested product and do not substitute other documents (e.g. administrative documents, certificates etc.) required by state supervising administration according to specific directives.
This report must not be reproduced without a written consent of VÚBP-ZL, partial reproduction is not allowed.

This test report was issued in Czech and English versions. Both versions have the same validity

| Phone | Web | E-mail | Bank connection | IČ: 00025950 |
|---|---|---|---|---|
| +420 221 015 811 | www.vubp.cz | | Praha 71336-011/0100 | DIČ: CZ00025950 |

### 1. Introduction – Basic Information

The tests were carried out on the basis of application No. S-058/2021 dated 11 February 2021.

Area of filter Phoenix is 900 cm2. Filter material consist of eight (8) layers of single-faced jersey fabric, double side fleece, 100% polyester. Mass per unit area of 1 layer (EN 12127): 162 g.cm-2. Number of columns per cm (EN 14971): 12, number of rows per cm: 12.Thickness of 1 layer under load 20±0,2g (EN ISO 5084): 2,28 mm.

The tests are intended for the needs of the client, sample of filter for laboratory tests was delivered by the client on 11 February 2021 in the number of 1 piece. Sample was registered in the Laboratory Register under number 643.

### 2. Indications of test specifications, methods and proceedings

The tests were performed according to the following standards:

EN 143:2000, EN 143:2000/A1:2006 Respiratory protective devices. Particle filters. Requirements, testing, marking

**Methods update**

Not used.

**Deviations and additions of test specifications**

The sample was tested in the as received condition.

### 3. Instruments used for tests

Hygrometer / barometer GFTB 200
Timer Ruhla
The apparatus for testing filters Sodium Chloride aerosol type MOORE'S 1100
NaCl aerosol generator type 4000

**Metrological support**

Metrological support pertaining to the devices is based on the VÚBP-ZL metrological regulations.

### 4. Tests

**Test results**

The tests were carried out in the respiratory protection laboratory of VÚBP-ZL.

**4.1 Filter penetration test** art. 8.7
Sodium Chloride aerosol test. Measured at 30 l/min and 95 l/min

*Initial penetration of sodium chloride aerosol*

| sample | conditioning | penetration in % 30 l/min | penetration in % 95 l/min |
|---|---|---|---|
| 643 | AR | 0,47 | 2,07 |

Notice: AR – As received

**Requirements for the penetration of filter material**

| Classification | Maximum penetration of test aerosols (%) | |
|---|---|---|
| | Sodium chloride test 95 l/min | Paraffin oil test 95 l/min |
| P1 | 20 | 20 |
| P2 | 6 | 6 |
| P3 | 0,05 | 0,05 |

Table of uncertainties of measurement

| Test number in report | Total expanded relative uncertainty in % |
|---|---|
| 4.1 | 4,47 |

These uncertainties of measurement are an expanded standard uncertainty calculated on the basis of the standard deviation multiplied by k = 2 (which guarantees a confidence interval of approximately 95%).

Photo No.1

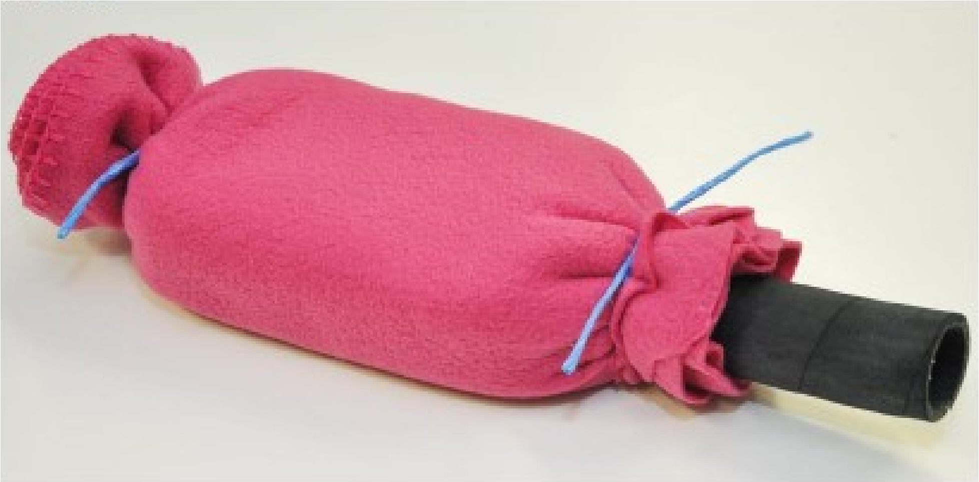

Test report elaborated by: Ing. Lukáš Zavřel

_________________________________end of Test report_________________________________

### 8. Pressure Drop of the Filter Cartridge

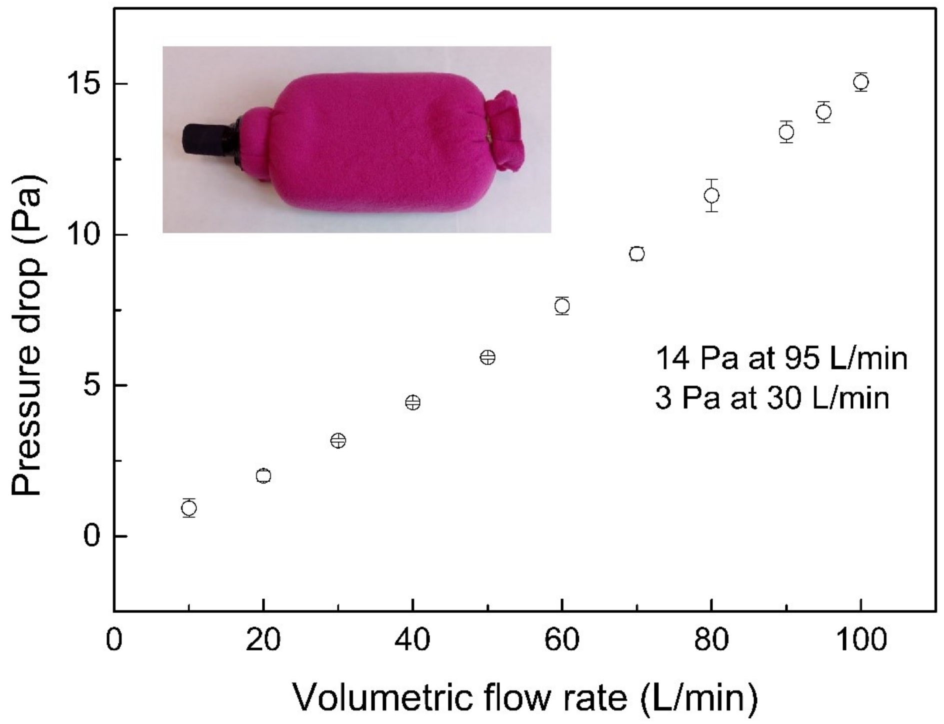

*Figure S8 shows the dependence between the pressure drop of the cartridge (8 layers of PES fabric, surface 900 cm$^2$) and the volumetric flow rate. Part of the pressure drop occurs in the hose (length 150 mm, inner diameter 30 mm) that is part of the cartridge. The low values of the pressure drop of the filter cartridge are explained by the relatively thick fibers of the PES fabric. The pressure drop is inversely proportional to the square of the fiber diameter.*[1] *The meltblown fibers used in typical respirators have a diameter of 2-7 µm*[2], *while the PES fabric is made of fibers with a diameter of 13 µm (Table S1).*

### 9. Comparison with FFRs (N95, KN95, and FFP2)

Our filter kit with a cartridge area of 900 $cm^2$ and 8 layers of polyester fleece kit reaches a filtration efficiency of 98 % and pressure drop of 84 Pa at 95 L/min. The corresponding Q factor is 20 $kPa^{-1}$ (the Q factor is explained in chapter 11). We compare our filter kit with N95-type filtering facepiece respirators (FFRs) (KN95 and FFP2), as this type is most commonly used in medical facilities.[3] Canadian studies from the end of 2020 compare the performance of 43 N95 FFRs (KN95 and FFP2) and provides the minimum filtration efficiency in the measurement range from 20 to 600 nm with a volumetric flow rate of 85 L/min.[4,5] The tested FFRs were installed on a support plate and sealed with adhesive tape. The penetration and pressure drop values are plotted for each FFR. The results are shown in Figure S9.

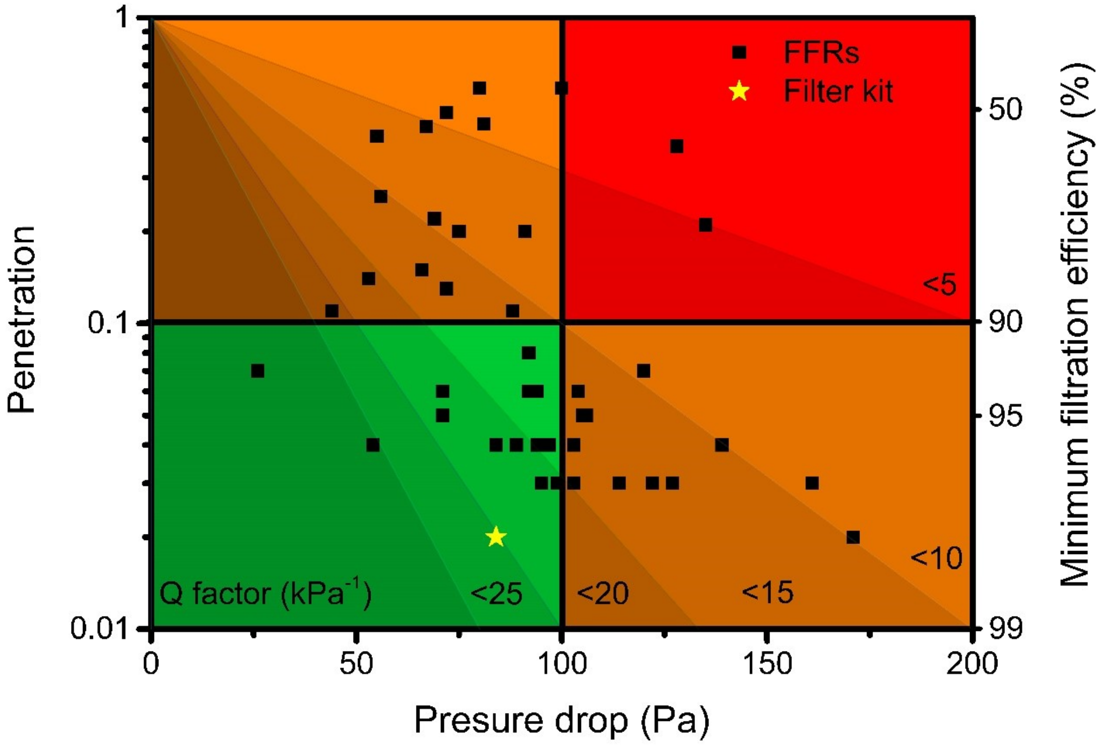

*Figure S9 shows the maximum penetration (minimum filtration efficiency) and pressure drop of 43 FFRs and the filtration kit at volumetric flow rates of 85 L/min and 95 L/min, respectively. The yellow star represents the performance of our filtration kit. The results are divided into four quadrants for easier orientation. The green quadrant contains FFRs with a filtration efficiency higher than 90 % and a pressure drop lower than 100 Pa. The six shaded areas show the Q factor values.*

### 10. *Filter Tunability up to 99.99 %*

Approximately 90 % of the pressure drop of the filter kit (84 Pa) is caused by the necessary accessories such as the hoses and half mask. Only 10 % of the pressure drop is caused by the polyester fleece knit filter material, i.e., 8 Pa in the case of eight layers and a filter area of 900 $cm^2$ = size 6 (see Figures 2 D and 4 in the original article). One layer of polyester fleece knit has a pressure drop of ca. 1 Pa. It follows that the filtration efficiency can be increased by increasing the number of layers with a minimal increase in the total pressure drop of the filter kit up to, for example, an efficiency of 99.9 % or 99.99 % (Figure S10, Figure S11). The calculation was performed on the basis of the evaluated coefficient of fractional penetration (Figure S15) and (Eq. S6).

For a filter with a filtration efficiency higher than 99 %, it would be expedient to use a full face mask that limits the amount of unfiltered bypassed air to 0.05 % rather than a half mask, where the quantity of unfiltered bypassed air can reach 1 %.

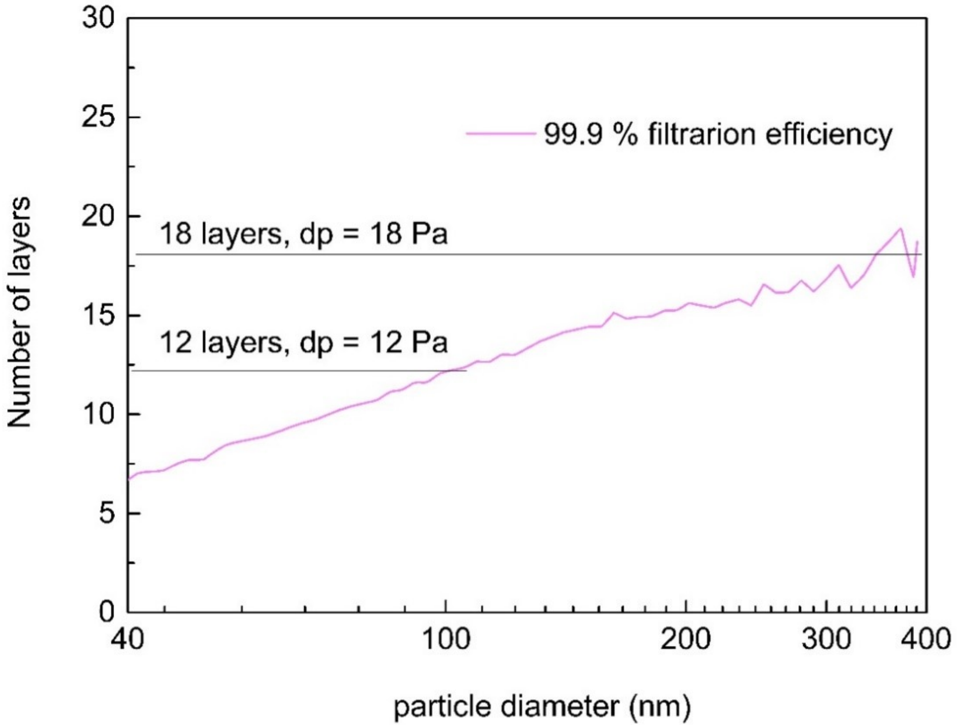

*Figure S10 shows the number of layers required to achieve a filtration efficiency of 99.9 % for a filter area of 900 $cm^2$ (size 6) depending on the diameter of the filtered particles. For particles with diameters of 100 and 350 nm, the number of layers and the corresponding pressure drops are marked.*

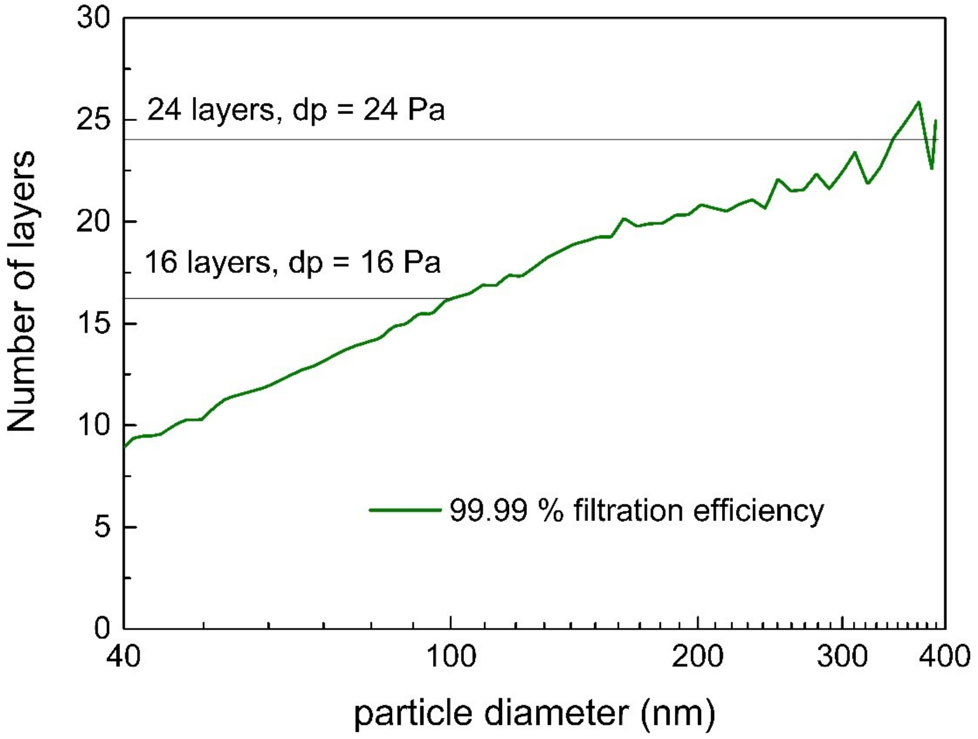

*Figure S11 shows the number of layers required to achieve a filtration efficiency of 99.99 % for a filter area of 900 cm$^2$ (size 6) depending on the diameter of the filtered particles. For particles with diameters of 100 and 350 nm, the number of layers and the corresponding pressure drops are marked.*

### 11. Quality Parameter Q of the Filter Fabrics

The filter quality $Q$ ($kPa^{-1}$) is a parameter combining the penetration $P$ and pressure drop $\Delta p$ of the filtering layer to determine the overall filter performance.

$$Q = \frac{log\frac{1}{P}}{\Delta p} \qquad \text{(Eq. S5)}$$

The filter quality is insensitive to the thickness of the filtering layer (in the first approximation) and thus useful for comparing different types of filtering materials. The dependence of the Q factor on the particle diameter is shown in Figure S12. The presented Q factor is the average of Q factors calculated from the size-resolved penetration measurements in Figure S6 (A), and the face velocity was 10.6 $cm.s^{-1}$. The knit polyester fleece fabric exhibits filter quality Q = 33 $kPa^{-1}$ for 100 nm particles. The most penetrating particle size (MPPS) in the case of PES knit fleece is a diameter of approximately 350 nm. The value of the Q factor for 350 nm particles is ca. 21 $kPa^{-1}$.

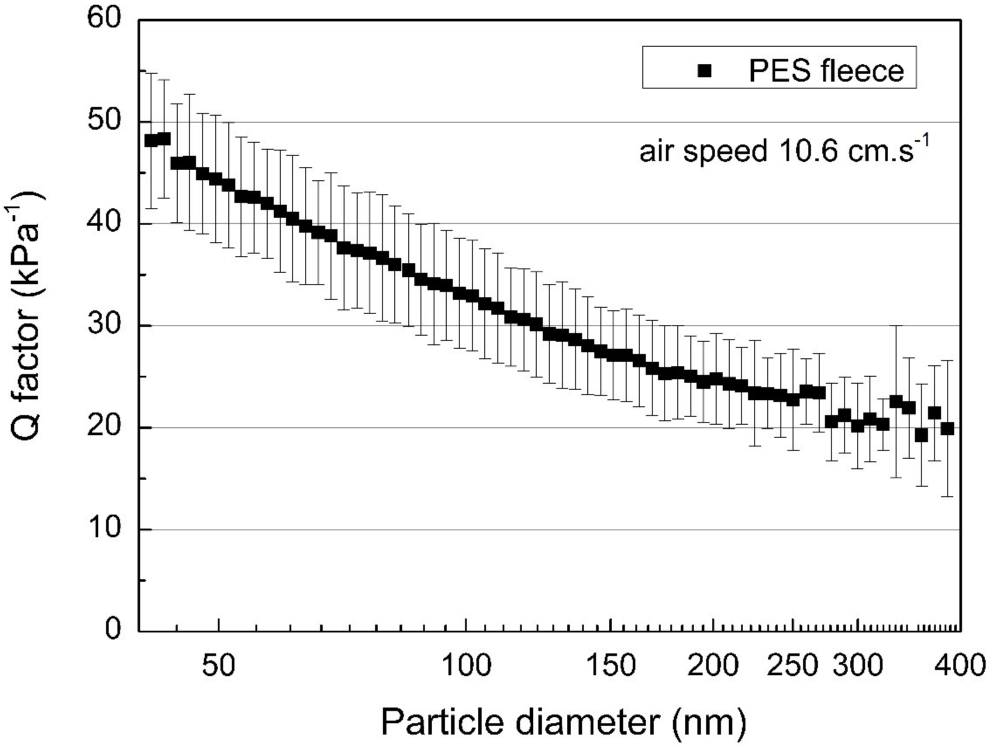

*Figure S12. Dependence of the Q factor of polyester fleece knit fabric on the particle diameter. The measurement was performed at a volumetric flow rate of 95 L/min, and the filter area corresponded to the size of a standard drape (150 $cm^2$).*

Recently, Zhao et al. tested household materials to determine their suitability for the manufacture of drapes.[6] Textile materials made of cotton, silk, polyester (without a fleece finish) and nonwoven polypropylene show a Q factor in the range from 3 to 17 $kPa^{-1}$. These are values lower than that of the polyester knit fleece that we tested. In addition, they were measured at a face velocity of 5.3 $cm.s^{-1}$. For this face velocity, the Q factor of the polyester knit fleece is equal to 47 $kPa^{-1}$. For use in the filtration of particulate matter, polyester knit fleece clearly outperforms other commonly available textile fabrics.[7,8] The reasons for this are explained in Figure S1 and Figure S8.

### 12. Filter Inlet on the Back → Reduced Inhaled Dose During AGMP

The superior performance of the filtration kit compared with that of an FFR can be seen in a situation where a person wearing our filter is in the proximity of a source of droplets containing virus particles, e.g., discussion with a patient, intubation, gastroscopy, or otorhinolaryngology (aerosol generating medical procedures).[9,10] We assume that a greater distance of the filter inlet from the virus source and its shielding by the body of the filter user results in a lower concentration of inhaled virus (Figure S13).

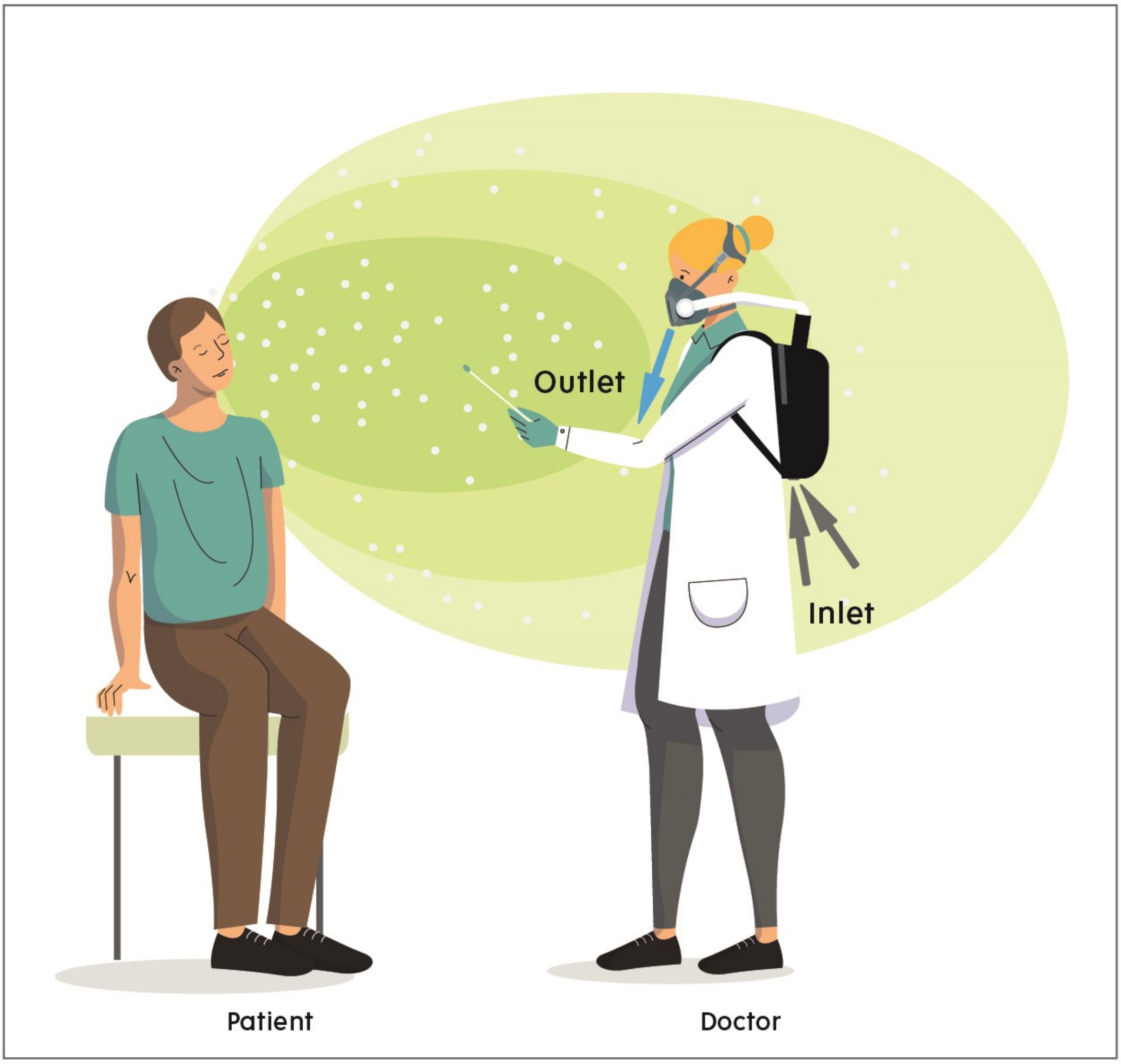

*Figure S13. The inhaled dose of aerosol emitted by the patient is decreased by the position of the filter inlet (in the case of a discussion or during aerosol generating procedures).*

### 13. The Price Of the Filter Kit and Its Components

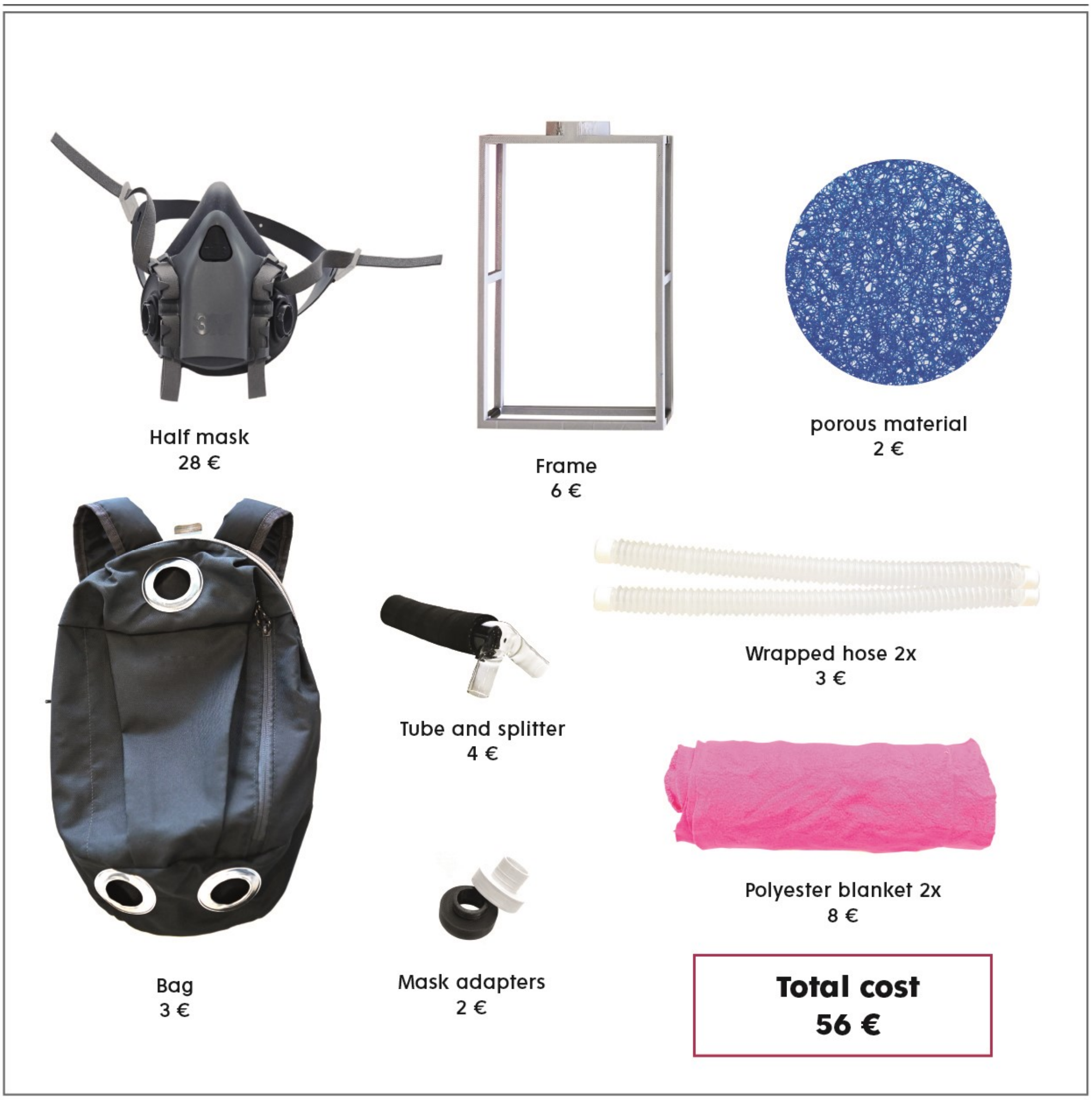

*Figure S14. The figure shows the components needed to assemble the filter kit including the retail prices. The half mask can be replaced with a less expensive model, and the 3D printed frame and porous material can be replaced with a simpler alternative (see Chapter 14).*

### 14. Supplementary Construction Details

Note that the presented filter kit can be assembled at home; however, it is recommended that a single manufacturer assemble the kit completely and thus guarantee the functionality of each individual part and at the same time provide instructions for regular maintenance and replacement of spare parts. The text below provides additional information about the individual components of the filter kit.

Filter material: PES fleece

In addition to the knitwear presented in the article, partial tests were performed on a knit PES fleece fabric sold by IKEA as a KRAKRIS blanket. The partial results of the filtration efficiency and pressure drop differed by less than 10 % from the results of the knit PES fleece fabric analyzed in the article. Two strips of the PES fabric measuring 160 x 40 cm were used to wrap the frame and create the filter cartridge. Before use, it is important to boil the PES fabric for ten minutes (preferred procedure) or wash it once at 95 °C.

Frame for cartridge assembly

The frame was printed using a 3D printer, as shown below, but alternatively, it can be made of hard paper or wood (see *Figure S2*). The thickness of the frame bars was 6 mm.

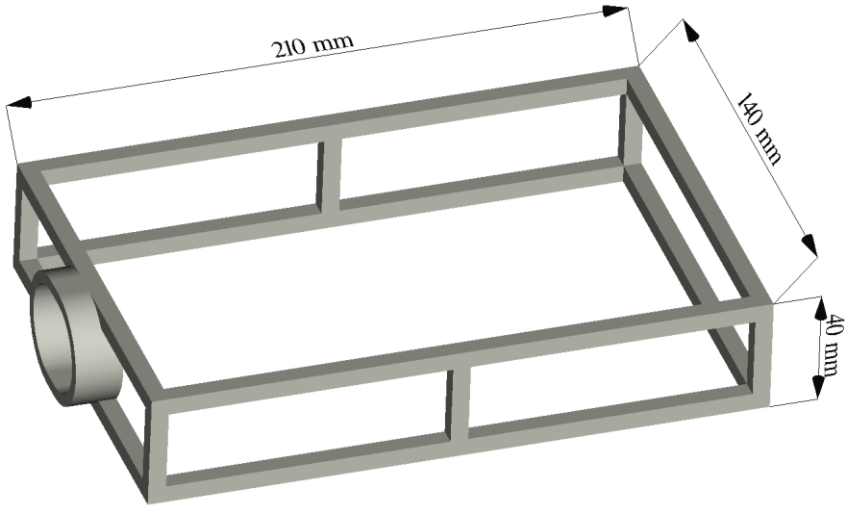

The filter cartridge with an area of 900 $cm^2$ and 8 layers of knit polyester fabric

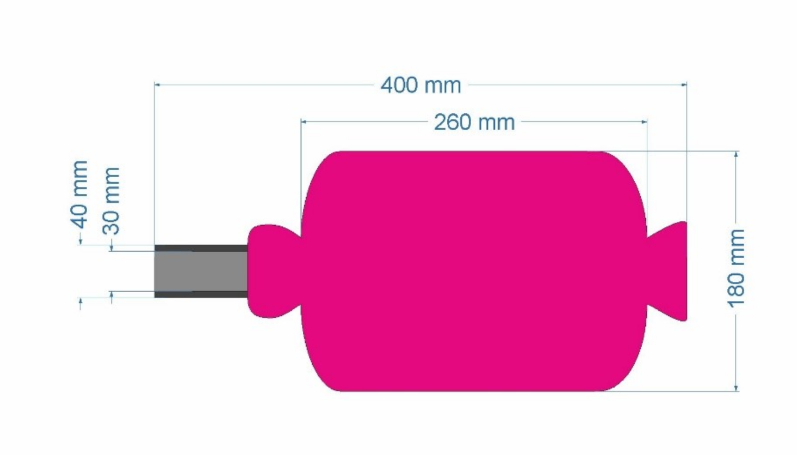

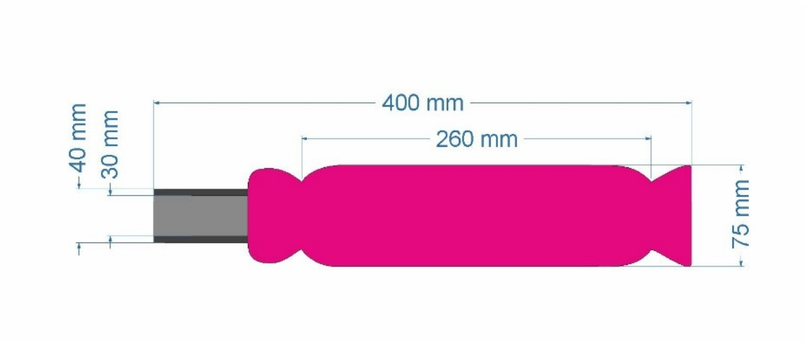

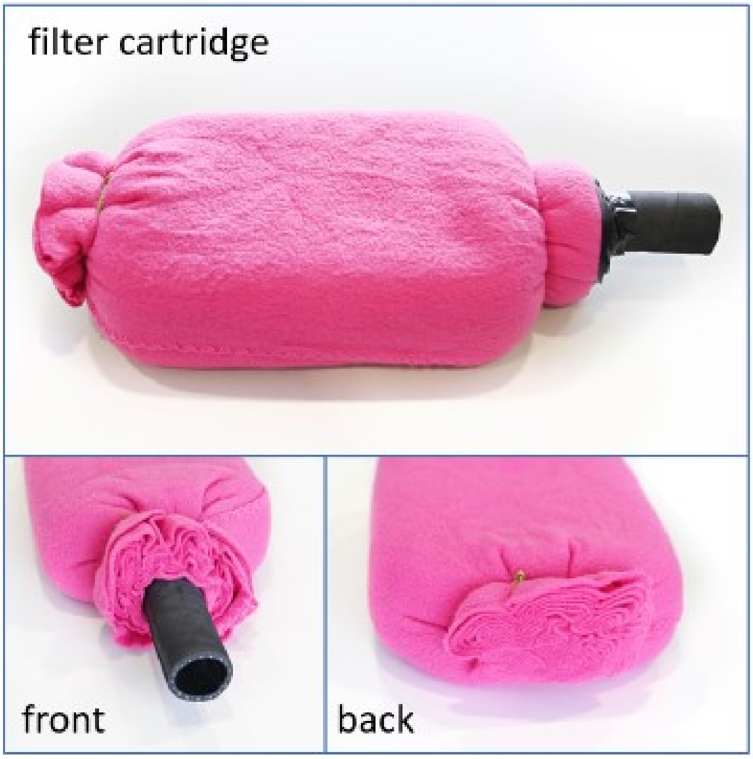

Air flow splitter

The splitter was made of glass, but plastic is a suitable alternative. It was designed so that it could be inserted inside a rubber hose and two flexible hoses with an inner diameter of 22 mm.

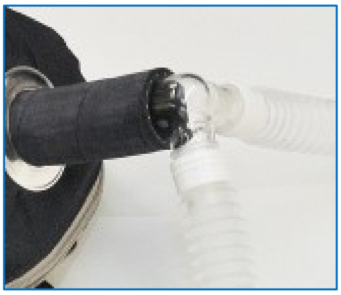

Adaptors

The adaptors were 3D printed to mimic the lock used for the 3M filter cartridge. To ensure a good seal of the joint, it is necessary to produce high-quality adaptors.

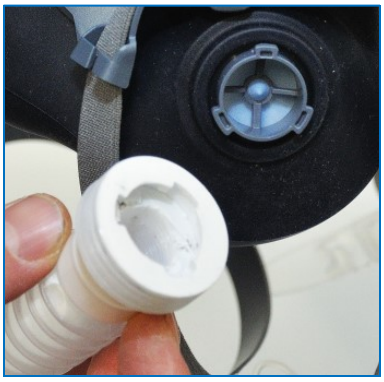

Porous material Matala FSM-365

The porous material Matala FSM-365 prevents the filter cartridge from coming into contact with the walls of the backpack so that it can pull in air over its entire surface. We chose a plate thickness of 20 mm for the porous material, which resulted in a minimal effect on the pressure drop of the kit. The porous material Matala FSM-365 can be replaced with spacers that define the space between the backpack and the filter cartridge to prevent the passage of air.

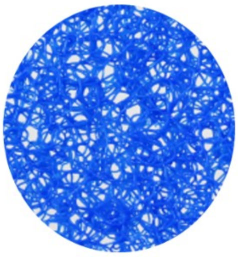

Backpack

The dimensions of the backpack must be such that they do not exert pressure on the filter cartridge. When the cartridge is under pressure, the polyester knit fleece fabric is compressed, resulting in an increased breathing resistance. A sufficient free space on each side of the backpack is ca. 10 mm.

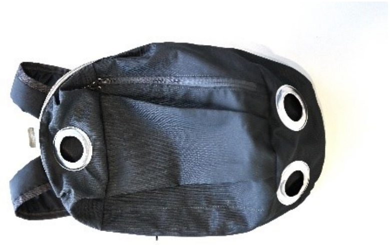

### 15. *Coefficient of Fractional Penetration*

To provide more detailed insight into the filter performance, the coefficient of fractional penetration γ is shown (Figure S15). Every single curve representing the coefficient of fractional penetration γ was calculated on the basis of the measured penetration for each filter size with 4, 6, and 8 layers (measurements with 1 and 2 layers were scattered and thus excluded). The penetration *P* can be expressed as a function of the filtering material thickness *L* (mm) and coefficient of fractional penetration γ ($mm^{-1}$):

$$P = \frac{1}{e^{\gamma L}} \qquad \textit{(Eq. S6)}$$

A higher value of γ means a lower value of penetration and thus better filtering performance. In our case, the coefficient of fractional penetration γ decreases while the particle size increases; a corresponding situation in the case of penetration is seen in Figure S15. A higher filter size (Table S2. Filter equivalent size) results in a higher coefficient of fractional penetration γ.

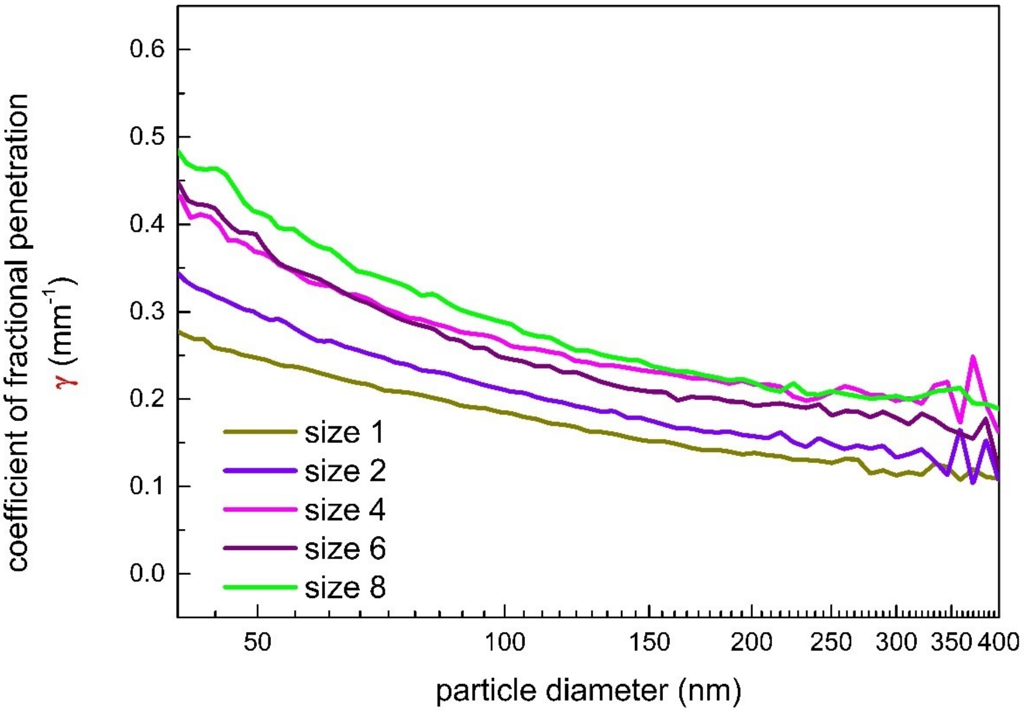

*Figure S15. Dependence of the coefficient of fractional penetration γ of polyester fleece knit fabric on particle diameter for different filter sizes.*

### *16. Particle Sized 1000 nm Captured In Polyester Fleece Knit Fabric*

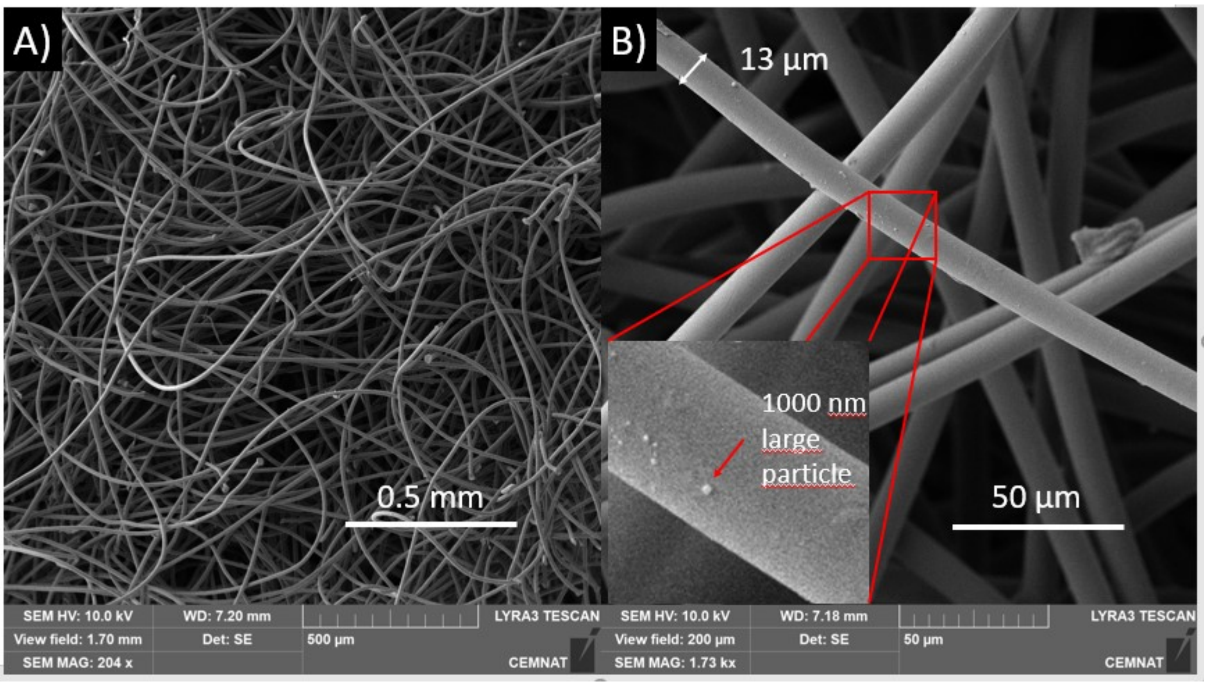

*Figure S16 shows a scanning electron microscopy image of a polyester knit fleece fabric. The fleece finish is visible, and the knit part of the fabric is deeper. In the enlarged section of picture B, there is a 1000 nm particle captured on the surface of a fiber. The virion of SARS-CoV-2 is 60-140 nm in diameter.*[11] *The average fiber thickness of polyester knit fleece fabric is 13 µm. The pore size is tens or hundreds of µm, and these values are much larger than those of nanofiber filters. Thus, clogging of the PES filter would be slower than that of nanofiber filters.*

### *17. References*

1. Hinds WC. Aerosol technology: properties, behaviour, and measurement of airborne particles. 2nd ed. Wiley-Interscience; 1999.
2. Drabek J, Zatloukal M. Meltblown technology for production of polymeric microfibers/nanofibers: A review. Phys Fluids. 2019;31. doi:10.1063/1.5116336
3. Frush K, Lee G, Wald SH, Hawn M, Krna C, Holubar M, et al. Navigating the Covid-19 Pandemic by Caring for Our Health Care Workforce as They Care for Our Patients. NEJM Catal. 2021;2: 1–34. doi:10.1056/cat.20.0378
4. Brochot C, Saidi MN, Bahloul A. How Effective Is the Filtration of 'KN95' Filtering Facepiece Respirators During the COVID-19 Pandemic? Ann Work Expo Heal. 2020; 1–9. doi:10.1093/annweh/wxaa101
5. Brochot C, Bahloul A. Qualitative Knowledge of Filtering Facepiece. J Int Soc Respir Prot. 2020;37, No. 2: 94–107.
6. Zhao M, Liao L, Xiao W, Yu X, Wang H, Wang Q, et al. Household Materials Selection for Homemade Cloth Face Coverings and Their Filtration Efficiency Enhancement with Triboelectric Charging. Nano Lett. 2020;20: 5544–5552. doi:10.1021/acs.nanolett.0c02211
7. Kwong LH, Wilson R, Kumar S, Crider YS, Reyes Sanchez Y, Rempel D, et al. Review of the Breathability and Filtration Efficiency of Common Household Materials for Face Masks. ACS Nano. 2021;15: 5904–5924. doi:10.1021/acsnano.0c10146
8. Zangmeister CD, Radney JG, Vicenzi EP, Weaver JL. Filtration Efficiencies of Nanoscale Aerosol by Cloth Mask Materials Used to Slow the Spread of SARS-CoV-2. ACS Nano. 2020;14: 9188–9200. doi:10.1021/acsnano.0c05025
9. Tran K, Cimon K, Severn M, Pessoa-Silva CL, Conly J. Aerosol generating procedures and risk of transmission of acute respiratory infections to healthcare workers: A systematic review. PLoS One. 2012;7. doi:10.1371/journal.pone.0035797
10. Lockhart SL, Duggan L V., Wax RS, Saad S, Grocott HP. Personal protective equipment (PPE) for both anesthesiologists and other airway managers: principles and practice during the COVID-19 pandemic. Can J Anesth. 2020;67: 1005–1015. doi:10.1007/s12630-020-01673-w
11. Zhu N, Zhang D, Wang W, Li X, Yang B, Song J, et al. A Novel Coronavirus from Patients with Pneumonia in China, 2019. N Engl J Med. 2020;382: 727–733. doi:10.1056/nejmoa2001017
